# Low back pain care pathways and costs: association with the type of initial contact health care provider. A retrospective cohort study

**DOI:** 10.1101/2022.06.17.22276443

**Authors:** David Elton, Thomas M. Kosloff, Meng Zhang, Protima Advani, Yinglong Guo, Scott T. Shimotsu, Sean Sy, Ari Feuer

**Author notes:** Corresponding author: David Elton, UnitedHealth Group, 5995 Opus Parkway, Minnetonka, MN 55343, Phone: -1349.

## Abstract

**Background:** Low back pain (LBP) is prevalent, management benefits from high-quality clinical practice guidelines, and yet LBP is a common source of low value care. The purpose of this retrospective cohort study was to examine the association between the type of initial contact health care provider (HCP), service utilization, and total episode cost for the management of LBP.

**Methods:** Episode of care was used to analyze a US national sample of LBP episodes completed in 2017-2019. A combined surgical and non-surgical (pooled) sample and a non-surgical sample were separately analyzed. The primary independent variable was the type of the initial contact HCP. Dependent measures included rate and timing of use of 14 types of health care services and total episode cost. The association between initial contact HCP, total episode cost and rate of prescription opioid and NSAID use was tested using a mixed effects model.

**Results:** The study included 616,766 continuously insured individuals aged 18 years and older with 756,631 episodes of LBP involving 386,795 HCPs and incurring $1,010,495,291 in expenditures. A primary care or specialist HCP was initially contacted in 62.0% of episodes, with these episodes associated with early use of low-value services such as imaging, pharmacologic, and interventional services. A non-prescribing HCP was initially contacted in 32.5% of episodes with these episodes associated with early use of guideline recommended first line services.

Each type of HCP emphasized different initial services with little indication of a stepped approach to managing LBP. Following adjustment for covariates chiropractors were associated with the lowest total episode cost. As an observational study of associations, numerous confounders may have impacted results.

**Conclusions:** An individual with LBP has different experiences based on the type of HCP initially contacted. Initial contact with primary care or specialist HCPs is associated with second- and third-line services provided before first line services, with little indication of a guideline recommended stepped approach to managing LBP. Increasing the likelihood of guideline-concordant, high-value care for LBP may require systemic changes to the health care delivery system. In the absence of red flags these changes may include increasing the proportion of individuals receiving early non-pharmacological treatment, either through improving direct access to non-prescribing HCPs or increasing timely referrals from primary care and specialist health care providers.

## Background

Health care policymakers, payors, professional societies, and other key stakeholders are seeking to implement strategies that improve the value of care for musculoskeletal disorders [1, 2]. Among musculoskeletal conditions, low back pain (LBP) causes the highest disease burden [1]. In the United States (US), 67% of individuals with LBP seek care annually [3], and LBP is the second most common reason for visiting a primary care provider [4]. In 2016 LBP and neck pain costs were estimated at $134 billion, collectively making them the most expensive medical disorders in the US, with commercial and public insurers funding more than 90% of spending [5].

In the absence of red flags indicating possible serious underlying pathology, LBP clinical practice guidelines (CPGs) emphasize self-care and non-pharmacologic interventions as a first-line management strategy [6–8]. Non-invasive and non-pharmacological physical treatments are viewed as cost-effective interventions [9] and are perceived to be of high-value for persons with LBP [10]. Care is characterized as low-value when individuals receive diagnostic testing (imaging) and treatment (opioids, spinal injections, and surgery) that are discordant with the recommendations of CPGs for LBP [11–13].

The gap between evidence-informed guidance and clinical practice results in overuse and misuse of low-value care, and underuse of high-value care [14]. Mismanagement of LBP has been estimated to be a source of almost half (46%) of low-value spending in US health care [15], and early exposure to non-guideline concordant care increases the risk of LBP transitioning from an acute to a chronic condition [16].

An evidence synthesis of studies published between 1995 and 2012 found for the treatment of acute/subacute LBP DCs had the highest percentage of concordance with CPGs (70.2%) followed by PTs (63.1%) and medical physicians (47.1%) [17]. Individual studies typically focused on a single type of health care provider (HCP) like; primary care physicians [18, 19], chiropractors (DCs) [20], physical therapists (PTs) [21–23], or surgeons [24]). Real-world guideline concordance could not be assessed in cross-sectional surveys that used clinical vignettes [20–23]. Retrospective cohort studies focused on specific services e.g., imaging utilization [18, 19] or multiple service outcomes but were not subdivided by initial HCP type [24].

The impact of the initial contact HCP has been employed as a method to evaluate variation in utilization and cost outcomes for LBP [25, 26]. Early access to DCs, PTs and licensed acupuncturists (LAcs) has been associated with lower rates of advanced spinal imaging studies [27–29] and lower rates of opioid prescriptions [26,30–32]. A recent study of lumbar surgical cases reported a low rate of early first-line conservative care was associated with high-cost and high-opioid use patterns [24].

The aim of this study was to examine the association between the type of initial contact HCP, total episodic service utilization and cost of care for the treatment of LBP in a US national sample of commercially insured adults. The hypothesis was that service utilization and total episode cost would vary based on the type of initial contact HCP.

## Methods

### Study design, population, setting and data sources

This is a retrospective cohort study of individuals seen by one or more HCPs for a complete episode of LBP. Multiple databases were linked to create a comprehensive database for analysis. The enrollee database included de-identified enrollment records, and administrative claims data for all inpatient and outpatient services, and pharmacy prescriptions, for enrollees from a single national commercial health insurer. The HCP database consisted of de-identified in and out-of-network HCP demographic information and professional licensure status. We extracted ZIP code level population race and ethnicity data from the US Census Bureau [33], adjusted gross income (AGI) data from the Internal Revenue Service [34], socioeconomic Area Deprivation Index (ADI) data from the University of Wisconsin Neighborhood Atlas^®^ database [35].

Due to the inability to control for important confounders such as patient preference for HCP type or specific services and lack of detailed understanding of the clinical complexity of an individual’s LBP, we elected not to control for typical confounders to generate incomplete causal insights [36, 37] through performing commonly used yet potentially inappropriate approaches such as propensity score matching [38]. As an alternative to blurring the line between association and causality through a process that simultaneously introduces distortion and complexity into results, actual measures of individual demographic attributes, episodic characteristics, and associations are provided for each type of HCP initially contacted by an individual with LBP.

Because data was linked from various sources, a review was performed to assess compliance with de-identification requirements. With data being de-identified or a Limited Data Set in compliance with the Health Insurance Portability and Accountability Act and customer requirements, the UnitedHealth Group Office of Human Research Affairs Institutional Review Board determined that this study was exempt from ethics review. The study was conducted and reported based on the Strengthening the Reporting of Observational Studies in Epidemiology (STROBE) guidelines (Supplement – STROBE Checklist) [39].

### Unit of analysis and cohort selection

Episode of care was selected as the unit of analysis. This approach has been shown to be a valid way to organize administrative claims data and develop a comprehensive profile of all services provided for a condition [40]. The *Symmetry^®^ Episode Treatment Groups^®^ (ETG^®^)* and *Episode Risk Groups^®^ (ERG^®^)* version 9.5 methodologies and definitions were used to translate administrative claims data into discrete episodes of care, which have been reported as a valid measurement for comparison of HCPs based on cost of care [41]. As it was possible for an individual to have multiple episodes of LBP during the study period, an episode sequence cohort categorization model was created (Figure 1) for the analysis.

**Figure 1.**
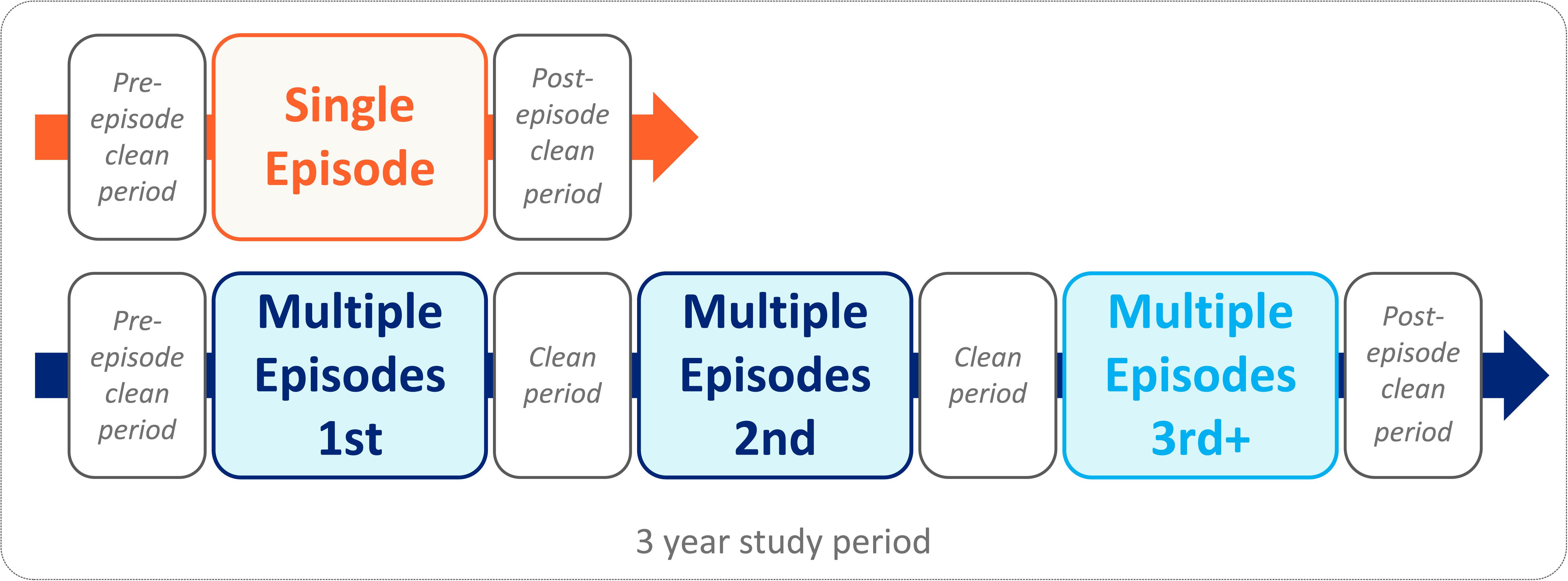
Episode sequence cohort and clean period conceptual model

Episodes with at least 91-day pre- and 61-day post-episode clean periods during which no services were provided by any HCP for any LBP diagnosis were included in the analysis. The episode duration was the number of days between the first and last date of service for each episode. The three-year study period, coupled with the 150-day pre and post episode clean periods, was associated with 0.31% of episodes with a duration of greater than 2 years. These episodes were excluded from the analysis. LBP episodes associated with diagnoses of malignant and non-malignant neoplasms, fractures and other spinal trauma, infection, congenital deformities and scoliosis, autoimmune disorders, osteoporosis, and advanced arthritis were also excluded from the analysis.

The cohort included individuals aged 18 years and older with a complete episode of LBP commencing and ending during the calendar years 2017-2019. This timeframe was selected to follow the release of the American College of Physicians (ACP) LBP CPG [6] in 2017 and before the influence of the COVID-19 epidemic on care patterns in early 2020. All individuals had continuous medical and pharmacy insurance coverage during the entire study period. Within the cohort two samples were created: a pooled sample consisting of episodes with and without a surgical procedure, and a non-surgical sample consisting of episodes without a surgical procedure. The pooled sample was created to comprehensively examine the patterns of initial contact HCP and subsequent management of LBP. Analysis of the non-surgical sample sought to partially address a study limitation of individuals of differing complexity selecting different types of initial contact HCPs (Supplement – Cohort).

### Variables

Data preprocessing, table generation, and initial analyses were performed in Python (*Python Language Reference, Version 3.7.5.*, n.d.). Linear mixed models regression was performed using R (*version 3.6.1*). A goodness-of-fit analysis was performed using D’Agostino’s K-squared test. Non-normally distributed data were reported using the median, interquartile range (IQR), quartile 1 (Q1), and quartile 3 (Q3).

The primary independent variable was the type of HCP initially contacted by an individual with LBP. Seventeen HCP types commonly contacted initially for an episode of LBP were analyzed. All HCP types could be accessed directly without a referral. Types of HCPs were segmented into primary care, non-prescribing, specialist, and emergency/urgent care HCP categories. Episodes initially contacting an HCP for whom a type could not be identified, often an out of network HCP, and a variety of non-physician HCP types infrequently initially contacted by individuals with LBP were excluded. Doctors of Osteopathy (DO) with evidence of billing an Osteopathic Manipulative Treatment Current Procedural Terminology*^®^* (CPT) code were separately reported, otherwise, DOs were included in the HCP type for which DOs were boarded. A Primary Care Provider (PCP) category was created consisting of Family Practice, Internal Medicine, General Medicine and OBGYN physician types. A Nurse category consisted primarily of nurse practitioners. An “MD-other” category was created for medical physician types not included in the other categories. For the 4.3% of episodes involving multiple, non-emergency/urgent care HCPs during the initial episode visit we assigned the initial type of HCP using the following hierarchy as we determined this to approximate the most likely experience of an individual with LBP:

a. Emergency Medicine/Urgent Care
b. Primary care
c. Non-prescriber
d. Specialist

The primary dependent variable was the rate and timing of use of 14 types of health care services segmented into first-, second-, and third-line service categories (Figure 2). The ACP CPG was used as the primary source to designate treatment interventions as first-, second-, or third-line [6].

**Figure 2.**
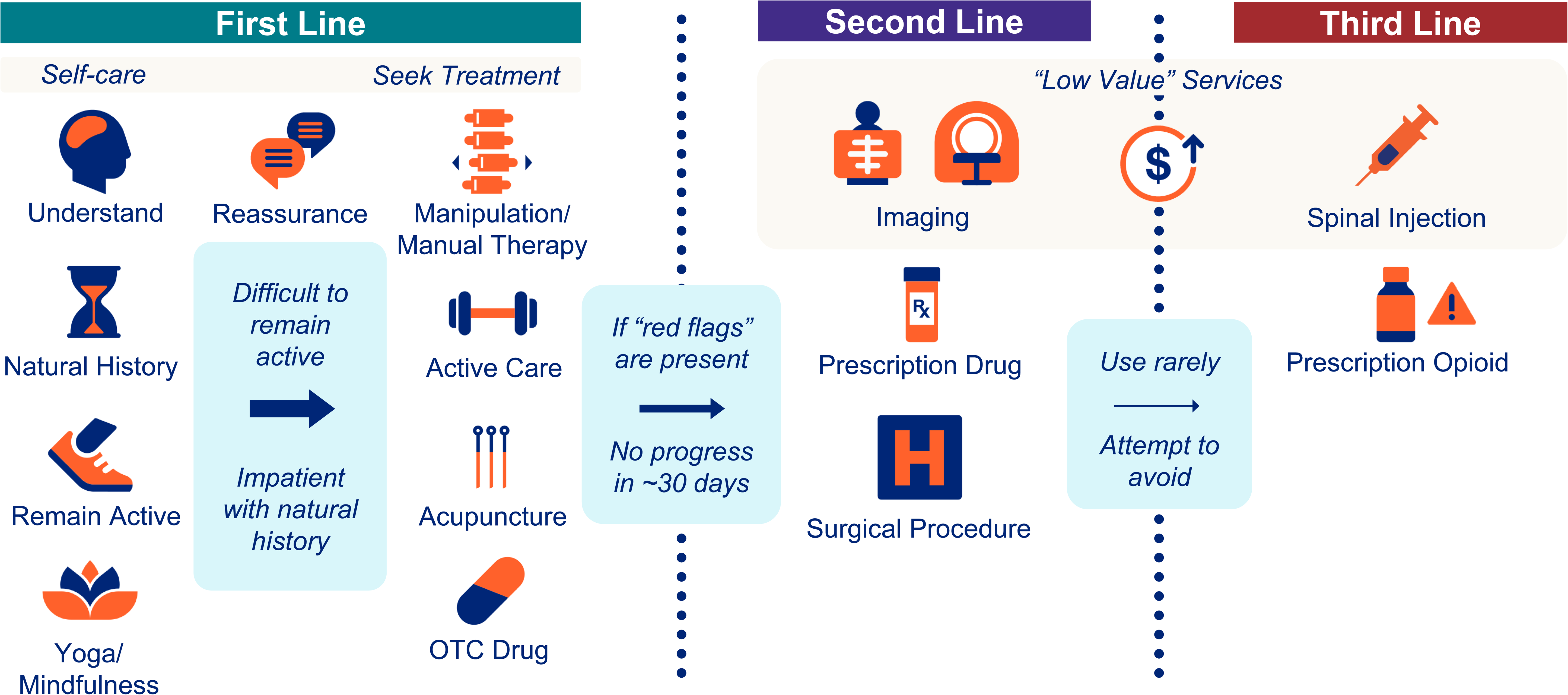
First-, second-, and third-line categorization of health care services reflecting clinical practice concordant stepped approach to managing low back pain

Odds (OR) and risk (RR) ratios, and associated 95% confidence intervals, were calculated for utilization of each service type for each type of HCP initially contacted by an individual with LBP. The baseline was episodes with a PCP as the initial contact HCP. Due to the tendency for ORs to exaggerate risk in situations where an outcome is relatively common, and as a measure more widely understood in associational analyses, RR were reported [42].

The secondary dependent variable was the total cost of care for all reimbursed services provided by any HCP during an episode. Total episode cost included costs associated with all services provided for LBP during an episode, including those not specifically identified in the 14 categories used in the analyses. Costs for services for which an insurance claim was not submitted, and indirect costs associated with missed days at work or reduced productivity, were not available.

Covariates included individual age, sex, and a comorbidity risk score (*Symmetry^®^ ERG^®^ version 9.5*, 2019). The risk score was included as a covariate as it evaluates an individual’s total all-cause illness burden in the measurement period.

The distribution of episodes among the episode sequence cohorts was summarized, pre- and post-episode clean periods and episode duration were calculated. For both the pooled and non-surgical samples, for each type of initial contact HCP we calculated the percent of episodes including each of the 14 types of health care service. Service utilization reflected services provided by any type of HCP an individual saw during the complete episode of LBP. The timing of when a service was first performed within an episode and total episode cost were calculated. Total episode cost was based on all services provided for LBP during an episode, including services (e.g., evaluation, durable medical equipment, lab studies, etc.) not included in the 14 categories of services described in the analysis.

For the bivariate analyses PCPs were selected as the reference group as PCPs were most frequently initially consulted for an episode of LBP. For bivariate analyses comparing results for each HCP type with the PCP reference group, Fischer’s Exact test (p value of .001) was used for comparing the percent of episodes including a service and Mann Whitney U test (p value of .001) was used for episode timing and total episode cost. A separate analysis was performed for each episode sequence cohort within the pooled and non-surgical samples.

For both pooled and non-surgical samples mixed effects regression models were used to test the relationship between type of initial contact HCP, total episode cost, rate of opioid use and rate of NSAID use. The reference group for mixed effects model was PCPs initially contacted by males of average cohort age with an *ERG^®^* score of zero. A unique identifier for the initial HCP was included as a random effect to account for individual HCP-influenced decision-making and cost differences.

## Results

The pooled sample included 616,766 individuals, with a median age of 45 (IQR 20), and 53.2% females. These individuals were associated with 756,631 complete LBP episodes involving 386,795 unique HCPs. For the pooled sample there were $1,010,495,291 in reimbursed health care expenditures a median total cost per episode of $208 (IQR Q1 $83, Q3 $660). The non-surgical sample included 600,390 individuals associated with 732,917 complete LBP episodes involving 356,316 unique HCPs. For the non-surgical sample there were $477,503,055 in reimbursed health care expenditures with a median total cost per episode of $196 (IQR Q1 $79, Q3 $577) (Table 1a). Individuals were from all 50 states and some U.S. territories (Supplement 1).

**Table 1a.**
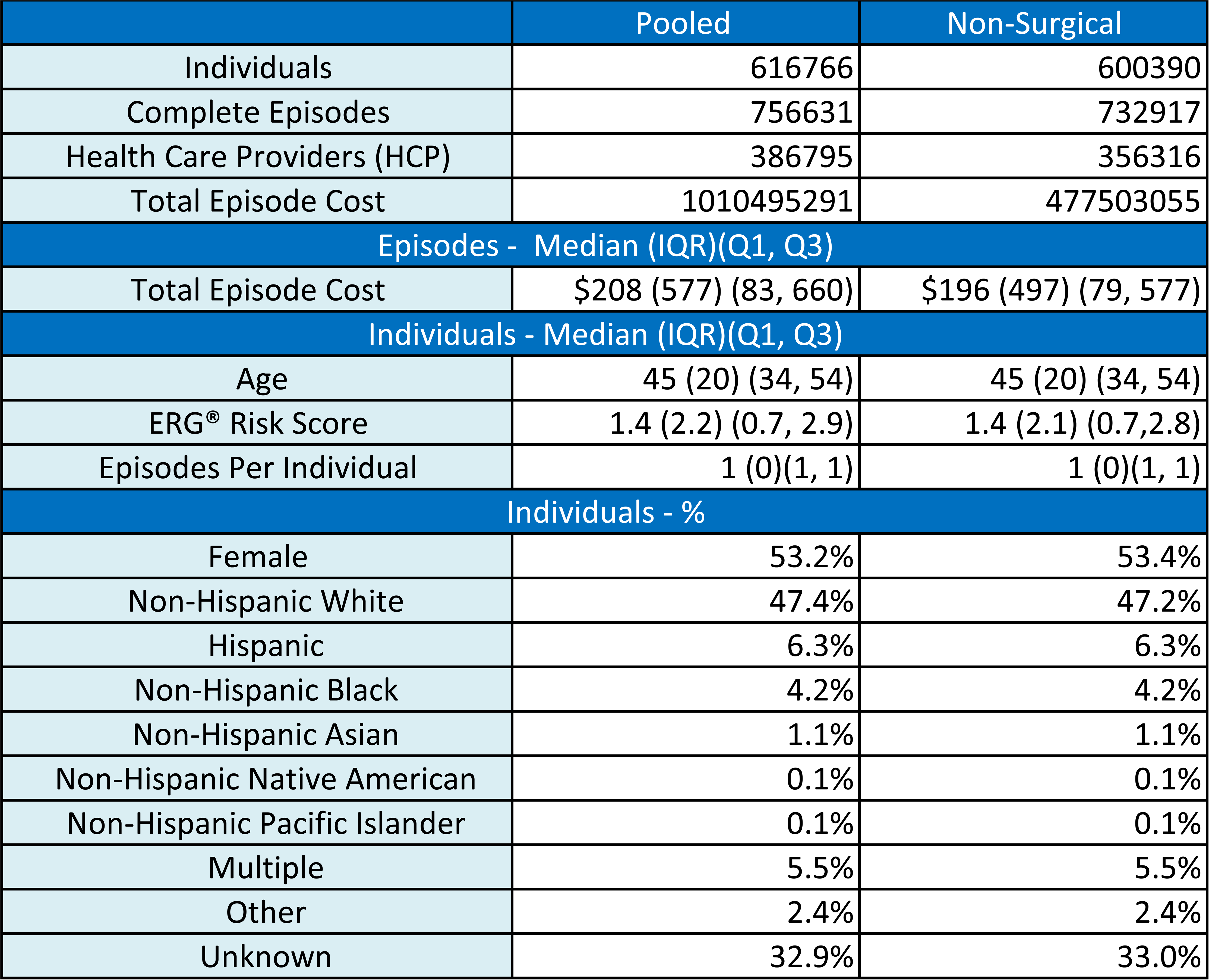
Low back pain cohort characteristics

|  | Pooled | Non-Surgical |
| --- | --- | --- |
| Individuals | 616766 | 600390 |
| Complete Episodes | 756631 | 732917 |
| Health Care Providers (HCP) | 386795 | 356316 |
| Total Episode Cost | 1010495291 | 477503055 |
| Episodes - Median (IQR)(Q1, Q3) |  |  |
| Total Episode Cost | \$208 (577) (83, 660) | \$196 (497) (79, 577) |
| Individuals - Median (IQR)(Q1, Q3) |  |  |
| Age | 45 (20) (34, 54) | 45 (20) (34, 54) |
| ERG® Risk Score | 1.4 (2.2) (0.7, 2.9) | 1.4 (2.1) (0.7,2.8) |
| Episodes Per Individual | 1 (0)(1, 1) | 1 (0)(1, 1) |
| Individuals - % |  |  |
| Female | 53.2% | 53.4% |
| Non-Hispanic White | 47.4% | 47.2% |
| Hispanic | 6.3% | 6.3% |
| Non-Hispanic Black | 4.2% | 4.2% |
| Non-Hispanic Asian | 1.1% | 1.1% |
| Non-Hispanic Native American | 0.1% | 0.1% |
| Non-Hispanic Pacific Islander | 0.1% | 0.1% |
| Multiple | 5.5% | 5.5% |
| Other | 2.4% | 2.4% |
| Unknown | 32.9% | 33.0% |

81.2% of individuals, generating 66.2% of episodes, had a single complete episode during the study period. The actual pre- and post-episode clean periods were substantially longer than the *ETG^®^* clean period definitions. For individuals with a single complete episode, the median pre-episode clean period was 640 days (IQR Q1 425, Q3 863). For individuals with multiple episodes, the median pre-episode clean period before the first of multiple episodes was 367 days (IQR Q1 200, Q3 578). The median number of days between sequential episodes was 196 (IQR Q1 118, Q3 326). The median post-episode clean period was 398 days (IQR Q1 236, Q3 607) (Supplement 2).

PCPs (35.7%) and DCs (31.0%) were the most common initial contact HCPs. Orthopedic surgeons (OS) (6.0%) were the most common initial contact specialist HCP. The characteristics of individuals, episodes and local population factors was variable for the different types of initial contact HCP. DC, LAc, and UC HCPs were initially contacted by younger individuals (median age approximately 41) and with a lower ERG® risk score (median approximately 1). LAc, PT, OS, PMR and UC HCPs were initially contacted by individuals from zip codes with lower levels of deprivation (ADI score less than 40) (Table 1b).

**Table 1b.** Type of health care provider (HCP) initially contacted by individuals with low back pain (LBP) and associated episode, individual and population characteristics

|  |  |  |  |  |  |  |  |  |
| --- | --- | --- | --- | --- | --- | --- | --- | --- |
| Median (IQR)(Q1, Q3) |  | Initial Contact HCP and Episodes |  |  |  | Individuals With LBP Initially Contacting HCP |  |  |
|  |  | Unique HCPs | Episodes | % of Episodes | Episodes per HCP | % Female | Age | ERG® Risk Score |
| Primary Care | PCP | 87557 | 269925 | 35.7% | 5 (7) (3, 10) | 53.3% | 47 (19) (37, 56) | 1.5 (2.3) (0.8, 3.0) |
|  | Nurse | 29074 | 53355 | 7.1% | 3 (4) (1, 5) | 56.7% | 45 (19) (35, 54) | 1.5 (2.3) (0.8, 3.0) |
|  | PA | 16881 | 34781 | 4.6% | 3 (4) (2, 6) | 53.0% | 45 (19) (35, 54) | 1.5 (2.3) (0.7, 3.0) |
|  | DO | 154 | 747 | 0.1% | 9 (16) (5, 21) | 66.1% | 47 (19) (36, 55) | 1.6 (2.2) (0.8, 3.0) |
| Non-Prescriber | DC | 23912 | 234868 | 31.0% | 20 (29) (10, 39) | 52.0% | 41 (19) (32, 51) | 1.0 (1.6) (0.5, 2.1) |
|  | PT | 5266 | 7143 | 0.9% | 2 (4) (1, 5) | 59.0% | 46 (20) (35, 55) | 1.9 (2.7) (0.9, 3.6) |
|  | LAc | 1880 | 4193 | 0.6% | 3 (6) (2, 8) | 65.5% | 41 (18) (33, 51) | 1.1 (2.1) (0.5, 2.6) |
| Specialist | OS | 11524 | 45363 | 6.0% | 9 (18) (3, 21) | 52.5% | 49 (20) (37, 57) | 2.2 (2.8) (1.0, 3.8) |
|  | PMR | 3773 | 19974 | 2.6% | 12 (16) (5, 21) | 53.5% | 49 (17) (39, 56) | 2.2 (2.9) (1.0, 3.9) |
|  | PM | 2577 | 7958 | 1.1% | 6 (8) (3, 11) | 53.1% | 50 (16) (41, 57) | 2.7 (3.3) (1.3, 4.7) |
|  | NS | 2478 | 7314 | 1.0% | 5 (7) (3, 10) | 48.4% | 51 (17) (41, 58) | 2.9 (3.4) (1.4, 4.8) |
|  | Neuro | 3870 | 7115 | 0.9% | 3 (3) (2, 5) | 62.9% | 50 (17) (40, 57) | 3.0 (3.8) (1.6, 5.4) |
|  | Rheum | 2511 | 6500 | 0.9% | 4 (7) (2, 9) | 73.3% | 51 (16) (42, 58) | 3.3 (3.5) (1.9, 5.4) |
|  | MD (Oth) | 13165 | 15070 | 2.0% | 1 (1) (1, 2) | 52.5% | 48 (20) (37, 57) | 2.3 (3.2) (1.2, 4.3) |
| Emergency/Urgent Care | EM | 10982 | 21545 | 2.8% | 3 (4) (1, 5) | 50.2% | 41 (20) (31, 51) | 1.4 (2.2) (0.6, 2.8) |
|  | Rad | 7845 | 15971 | 2.1% | 3 (5) (1, 6) | 57.3% | 47 (21) (35, 56) | 1.9 (3.0) (0.9, 3.9) |
|  | UC | 1286 | 4809 | 0.6% | 6 (11) (3, 14) | 51.2% | 41 (19) (32, 51) | 1.0 (1.7) (0.4, 2.1) |

| Median (IQR)(Q1, Q3) |  | Episode Attributes |  |  |  | Individual With LBP Home Address Zip Code Population Attributes |  |  |
| --- | --- | --- | --- | --- | --- | --- | --- | --- |
|  |  | # Different HCP Seen During Episode | Episode Duration | Pre-episode Clean Period | Post-episode Clean Period | Population % Non-Hispanic White (NHW) | Area Deprivation Index (ADI) | Household Adjusted Group Income (AGI) |
| Primary Care | PCP | 2 (2) (1, 3) | 28 (159) (1, 160) | 601 (456) (387, 843) | 436 (421) (277, 698) | 71% (36%) (48%, 85%) | 46.9 (34.0) (29.8, 63.7) | 62505 (37569) (48941, 86510) |
|  | Nurse | 2 (2) (1, 3) | 26 (154) (1, 155) | 643 (433) (437, 870) | 407 (400) (245, 645) | 77% (31%) (58%, 89%) | 55.0 (30.7) (38.8, 69.4) | 56647 (27050) (46750, 73801) |
|  | PA | 2 (3) (1, 4) | 22 (140) (1, 141) | 642 (440) (431, 871) | 417 (408) (251, 659) | 76% (30%) (57%, 87%) | 47.8 (31.2) (32.5, 63.6) | 61668 (32692) (49591, 82283) |
|  | DO | 1 (1) (1, 2) | 29 (154) (1, 156) | 593 (456) (387, 843) | 445 (413) (286, 699) | 77% (24%) (63%, 87%) | 40.4 (31.7) (26.0, 57.7) | 72779 (52091) (55162, 107254) |
| Non-Prescriber | DC | 1 (1) (1, 2) | 29 (115) (3, 118) | 634 (430) (429, 859) | 434 (423) (257, 680) | 78% (27%) (61%, 89%) | 43.6 (31.8) (28.0, 59.8) | 67641 (39020) (53607, 92627) |
|  | PT | 3 (2) (2, 4) | 62 (156) (26, 182) | 585 (439) (390, 829) | 432 (412) (272, 684) | 73% (30%) (55%, 84%) | 33.3 (32.7) (19.0, 51.7) | 78111 (58051) (56802, 114853) |
|  | LAc | 1 (1) (1, 2) | 34 (81) (7, 88) | 653 (436) (429, 865) | 446 (430) (253, 683) | 61% (36%) (40%, 76%) | 23.1 (24.3) (12.6, 37.0) | 90081 (66620) (64474, 131095) |
| Specialist | OS | 2 (3) (1, 4) | 43 (159) (1, 160) | 584 (477) (361, 838) | 449 (448) (280, 728) | 70% (33%) (50%, 83%) | 38.8 (35.1) (22.0, 57.2) | 73422 (55080) (54163, 109243) |
|  | PMR | 3 (3) (1, 4) | 74 (219) (10, 229) | 528 (543) (278, 821) | 433 (423) (308, 731) | 71% (32%) (52%, 83%) | 36.1 (35.1) (19.8, 54.9) | 75836 (60071) (55321, 115393) |
|  | PM | 2 (3) (1, 4) | 85 (254) (9, 263) | 515 (553) (256, 809) | 412 (368) (328, 696) | 70% (32%) (50%, 83%) | 45.4 (32.1) (28.7, 60.9) | 65469 (41582) (50822, 92403) |
|  | NS | 4 (5) (2, 7) | 104 (222) (20, 242) | 506 (573) (223, 796) | 453 (434) (319, 753) | 74% (31%) (54%, 86%) | 44.9 (34.6) (28.3, 62.8) | 67372 (44415) (51068, 95483) |
|  | Neuro | 3 (3) (1, 4) | 104 (229) (15, 244) | 521 (513) (282, 795) | 433 (407) (313, 720) | 68% (35%) (47%, 82%) | 41.8 (35.0) (24.7, 59.7) | 67895 (46558) (51440, 97998) |
|  | Rheum | 2 (3) (1, 4) | 105 (246) (7, 253) | 534 (516) (284, 800) | 425 (388) (316, 704) | 70% (35%) (49%, 84%) | 44.8 (34.7) (27.4, 62.1) | 65144 (43154) (49995, 93148) |
|  | MD Oth | 2 (2) (1, 3) | 67 (193) (1, 194) | 590 (454) (381, 835) | 426 (405) (271, 676) | 71% (34%) (50%, 84%) | 42.4 (34.6) (25.8, 60.4) | 68181 (45530) (51669, 97199) |
| Emergency/Ur gent Care | EM | 3 (2) (2, 4) | 2 (61) (1, 62) | 671 (441) (447, 888) | 445 (436) (256, 692) | 70% (41%) (44%, 85%) | 50.3 (33.7) (32.9, 66.6) | 58426 (32692) (46208, 78899) |
|  | Rad | 2 (1) (1, 2) | 1 (0) (1, 1) | 707 (444) (468, 912) | 446 (439) (255, 694) | 73% (37%) (50%, 87%) | 46.6 (35.1) (28.9, 64.0) | 61755 (38053) (48573, 86626) |
|  | UC | 2 (2) (1, 3) | 1 (30) (1, 31) | 689 (438) (458, 896) | 445 (435) (256, 691) | 68% (38%) (46%, 84%) | 38.1 (31.0) (23.5, 54.5) | 67990 (40923) (51750, 92673) |

PCP=Primary Care Provider, PA=Physician Assistant, DO=Doctor of Osteopathy, DC=Doctor of Chiropractic, PT=Physical Therapist, LAc=Licensed Acupuncturist, OS=Orthpedic Surgeon, PMR=Physical Medicine & Rehabilitation, PM=Pain Management, NS=Neurosurgeon, Neuro=Neurologist, Rheum=Rheumatologist, MD Oth=Other MD specialty, EM=Emergency Medicine, Rad=Radiologist, UC=Urgent Care, IQR=Interquartile Range

The most frequently provided first-line services were chiropractic manipulation (33.5% of episodes), active care (19.3%), and passive therapy (15.4%). The most frequently provided second-line services were radiographs (25.6%) and prescription NSAIDs (23.1%). The most frequently provided third-line services were opioids (16.2%) and spinal injections (6.7%). 3.1% of episodes included spinal surgery. Among first- and second-line services there was little indication of a stepped approach to managing LBP. If a service was provided, other than MRI, the median days into the episode when initially provided was within 7 days, and often on the initial visit. (Table 1c).

**Table 1c.** Services provded for low back pain

| Median (IQR)(Q1, Q3) |  | Episodes Including Service | % of Episodes | Days Into Episode When Initially Provided |
| --- | --- | --- | --- | --- |
| First Line | Manipulation - Chiropractic | 253643 | 33.5% | 0 (0) (0, 0) |
|  | Active Care | 146182 | 19.3% | 6 (35) (0, 35) |
|  | Passive Therapy | 116843 | 15.4% | 0 (19) (0, 19) |
|  | Manual Therapy | 97635 | 12.9% | 7 (42) (0, 42) |
|  | Acupuncture | 6870 | 0.9% | 0 (24) (0, 24) |
|  | Manipulation - Osteopathic | 6579 | 0.9% | 0 (12) (0, 12) |
| Second Line | Imaging - Radiography | 193767 | 25.6% | 0 (20) (0, 20) |
|  | Rx - NSAID | 175155 | 23.1% | 0 (33) (0, 33) |
|  | Rx - Skeletal Muscle Relaxant | 165386 | 21.9% | 0 (14) (0, 14) |
|  | Imaging - MRI | 77837 | 10.3% | 22 (79) (5, 84) |
| Third Line | Rx - Opioid | 122672 | 16.2% | 2 (58) (0, 58) |
|  | Spinal Injection | 50689 | 6.7% | 42 (100) (10, 110) |
|  | Spinal Surgery | 23714 | 3.1% | 44 (93) (14, 107) |
|  | Imaging - CT | 13672 | 1.8% | 16 (103) (0, 103) |

The percent of episodes including first-, second- and third-line services, and the timing of when these services were first introduced during an episode, was variable among types of HCPs initially contacted by individuals with LBP. For each episode sequence cohort within both pooled and non-surgical samples, initial contact with primary care, specialist and emergency/urgent care HCPs was associated with second- and third-line services provided most often, and if provided, typically within the first 7 days of an episode and commonly during the initial visit. Initial contact with non-prescribing HCPs was associated with one or more first-line therapies provided during the initial visit. The type of first-line therapy was variable for each type of non-prescribing HCP. Second- and third-line services, other than radiography, are infrequently provided to individuals with LBP initially contacting a non-prescribing HCP and if provided, are introduced later in an episode. Tables 2a and 2b present these data for the non-surgical sample and single episode cohort. For the most common HCPs initially contacted by an individual with LBP Figure 3 compares the different patterns in service use. Supplement – Care Pathways illustrates these patterns for additional types of HCP. For the non-surgical sample Figure 4 presents the RR comparing each type of initial contact HCP with the PCP reference.

**Figure 3.**
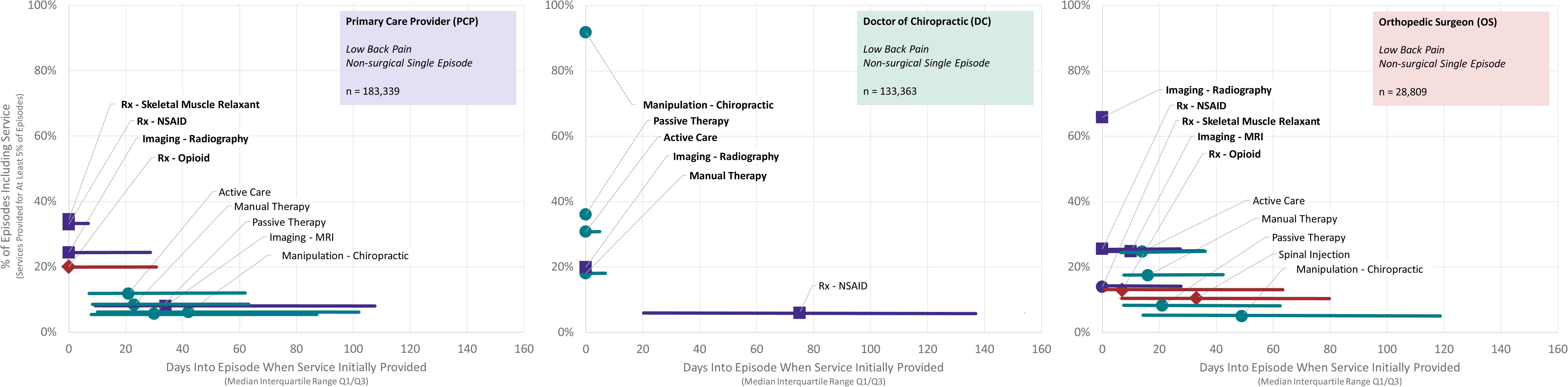
Rate and timing of use of health care services for individuals with low back pain initially contacting a primary care provider, chiropractor, and orthopedic surgeon - non-surgical single episode cohort

**Figure 4.**
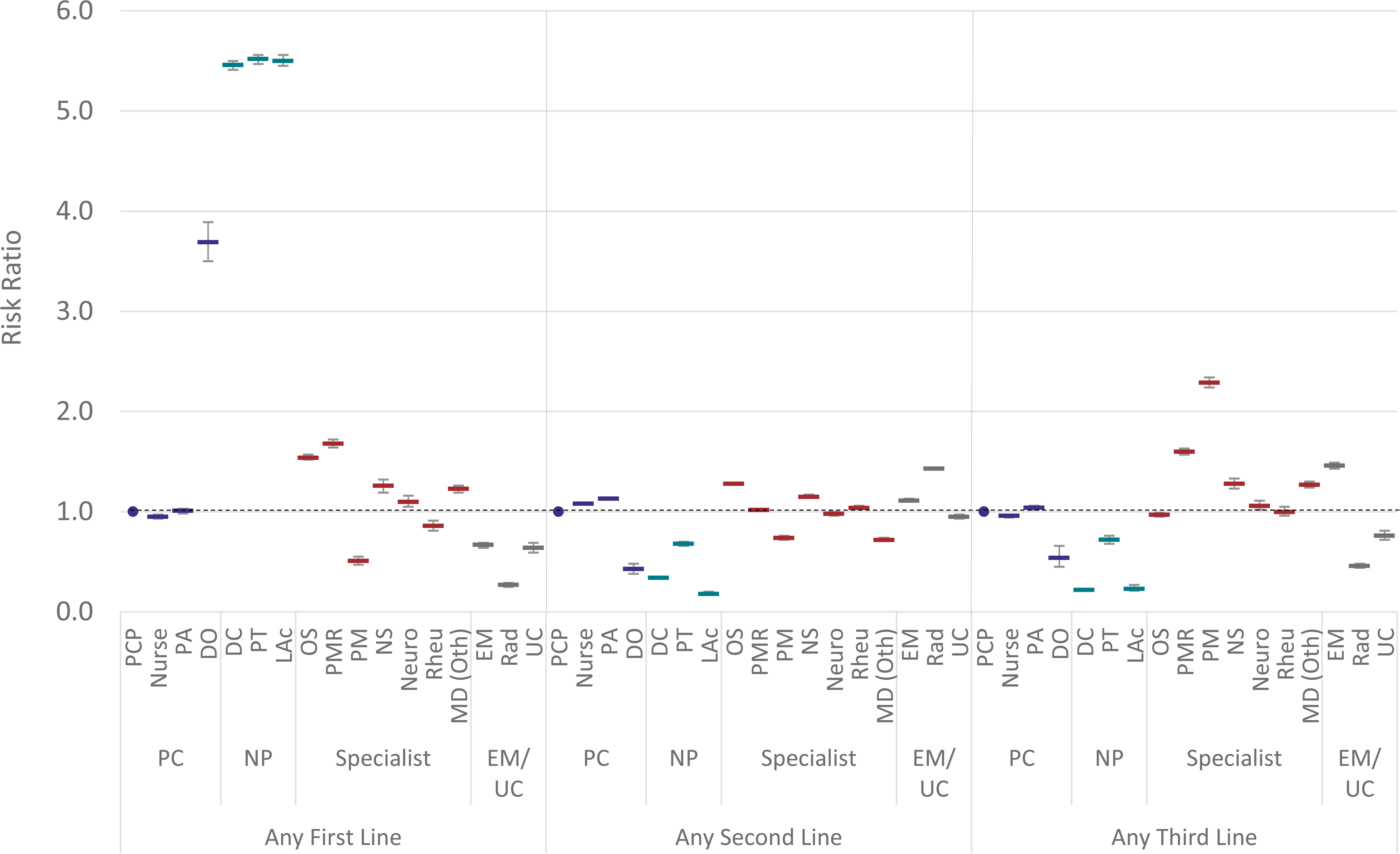
Risk ratio and 95% confidence interval for non-surgical low back pain episodic service exposure by initial contact health care provider compared to PCP reference.PC=Primary Care, NP=Non-Prescriber, EM/UC=Emergency Medicine/Urgent Care, PCP=Primary Care Provider; PA=Physician Assistant; DO=Doctor of Osteopathy; DC=Doctor of Chiropractic; PT=Physical Therapist; LAc=Licensed Acupuncturist; OS=Orthopedic Surgeon; PMR=Physical Medicine and Rehabilitation; PM=Pain Management, NS=Neurosurgeon; Neuro=Neurologist; Rheum=Rheumatologist; MD (Oth)=Other medical physician specialties, EM=Emergency Medicine; Rad=Radiologist; UC=Urgent Care

**Table 2a.** Low back pain % of episodes including service by type of initial contact health care provider (HCP)

|  |  |  |  |  |  |  |  |  |  |  |  |  |  |  |  |  |  |  |  |  |
| --- | --- | --- | --- | --- | --- | --- | --- | --- | --- | --- | --- | --- | --- | --- | --- | --- | --- | --- | --- | --- |
| Non-Surgical Single Episode Cohort |  | Episodes |  | First Line |  |  |  |  |  |  | Second Line |  |  |  |  | Third Line |  |  |  |  |
|  |  | Count | % | Any | Manipulation - Chiropractic | Active Care | Passive Therapy | Manual Therapy | Acupuncture | Manipulation - Osteopathic | Any | Imaging - Radiography | Rx - NSAID | Rx - Skeletal Muscle Relaxant | Imaging - MRI | Any | Rx-Opioid | Spinal Injection | Spinal Surgery | Imaging-CT |
| Total |  | 485388 | 100.0% | 40.6% | 29.6% | 18.7% | 14.5% | 12.4% | 0.9% | 0.8% | 56.8% | 27.9% | 23.5% | 23.5% | 8.9% | 18.8% | 14.9% | 4.5% | 0.0% | 1.6% |
| Primary Care | PCP-reference | 183339 | 35.3% | 17.4% | 6.1% | 11.8% | 5.6% | 8.4% | 0.3% | 1.5% | 65.6% | 24.4% | 33.0% | 34.5% | 8.0% | 22.8% | 20.0% | 3.8% | 0.0% | 1.3% |
|  | Nurse | 36891 | 7.1% | 16.3% | 7.4% | 11.0% | 5.7% | 7.1% | 0.1% | 0.4% | 70.5% | 25.0% | 35.3% | 41.0% | 7.4% | 21.9% | 18.2% | 4.7% | 0.0% | 1.6% |
|  | PA | 24302 | 4.7% | 17.4% | 6.5% | 13.1% | 5.0% | 8.8% | 0.3% | 0.3% | 73.8% | 30.5% | 33.7% | 40.5% | 9.7% | 24.1% | 19.6% | 5.8% | 0.0% | 1.8% |
|  | DO | 470 | 0.1% | 62.3% | 4.5% | 16.4% | 5.1% | 9.1% | 0.9% | 53.4% | 28.5% | 11.5% | 10.9% | 11.7% | 7.0% | 15.1% | 10.0% | 6.8% | 0.0% | 0.9% |
|  | All | 245002 | 47.2% | 17.3% | 6.3% | 11.8% | 5.6% | 8.2% | 0.3% | 1.3% | 67.1% | 25.1% | 33.4% | 36.0% | 8.1% | 22.8% | 19.7% | 4.1% | 0.0% | 1.4% |
| Non-Prescriber | DC | 133363 | 25.7% | 96.0% | 91.7% | 30.8% | 36.1% | 18.1% | 0.6% | 0.2% | 26.7% | 20.0% | 5.9% | 4.2% | 2.5% | 5.8% | 4.5% | 1.3% | 0.0% | 0.3% |
|  | PT | 4894 | 0.9% | 98.0% | 9.6% | 95.6% | 26.3% | 74.0% | 1.4% | 0.6% | 43.7% | 24.4% | 12.7% | 10.1% | 17.7% | 16.7% | 9.1% | 9.5% | 0.0% | 1.4% |
|  | LAc | 2642 | 0.5% | 97.8% | 8.6% | 17.9% | 41.2% | 44.0% | 88.9% | 0.4% | 11.8% | 5.2% | 5.0% | 2.5% | 3.2% | 5.8% | 4.2% | 1.7% | 0.0% | 0.3% |
|  | All | 140899 | 27.2% | 96.1% | 87.3% | 32.8% | 35.9% | 20.5% | 2.3% | 0.2% | 27.0% | 19.9% | 6.2% | 4.4% | 3.1% | 6.1% | 4.7% | 1.6% | 0.0% | 0.4% |
| Specialist | OS | 28809 | 5.6% | 28.0% | 5.0% | 24.7% | 8.2% | 17.4% | 0.3% | 0.6% | 83.3% | 65.8% | 25.5% | 13.9% | 24.8% | 21.7% | 13.0% | 10.5% | 0.0% | 2.0% |
|  | PMR | 12238 | 2.4% | 31.1% | 5.3% | 27.4% | 9.0% | 19.6% | 1.0% | 1.4% | 67.8% | 38.2% | 23.5% | 17.2% | 24.8% | 35.8% | 19.7% | 21.5% | 0.0% | 1.5% |
|  | PM | 4445 | 0.9% | 9.4% | 1.4% | 8.0% | 2.9% | 5.4% | 0.0% | 0.7% | 50.1% | 15.8% | 17.0% | 18.5% | 18.6% | 53.8% | 33.1% | 28.7% | 0.0% | 1.3% |
|  | NS | 3450 | 0.7% | 22.8% | 4.7% | 19.7% | 7.3% | 14.2% | 0.2% | 0.2% | 74.5% | 39.4% | 18.6% | 16.6% | 43.4% | 30.9% | 17.1% | 14.8% | 0.0% | 7.7% |
|  | Neuro | 4127 | 0.8% | 20.5% | 7.2% | 15.6% | 6.7% | 10.4% | 0.3% | 0.4% | 65.4% | 17.0% | 23.9% | 26.1% | 30.3% | 26.2% | 18.0% | 10.2% | 0.0% | 2.6% |
|  | Rheu | 3690 | 0.7% | 16.2% | 7.5% | 10.8% | 5.3% | 7.7% | 0.5% | 0.2% | 70.9% | 41.3% | 29.5% | 18.0% | 13.8% | 23.0% | 17.1% | 7.8% | 0.0% | 1.5% |
|  | MD (Oth) | 9385 | 1.8% | 21.9% | 12.6% | 11.7% | 7.4% | 8.1% | 0.8% | 1.2% | 49.6% | 21.1% | 24.5% | 17.9% | 9.5% | 31.4% | 27.6% | 4.9% | 0.0% | 2.1% |
|  | All | 66144 | 12.8% | 25.1% | 6.2% | 20.7% | 7.6% | 14.5% | 0.5% | 0.8% | 71.1% | 45.2% | 24.2% | 16.5% | 22.9% | 28.7% | 18.4% | 13.0% | 0.0% | 2.2% |
| Emergency/ Urgent Care | EM | 16993 | 3.3% | 10.9% | 4.1% | 7.8% | 3.9% | 5.4% | 0.2% | 0.3% | 71.4% | 32.2% | 36.1% | 41.9% | 6.6% | 34.4% | 27.5% | 2.4% | 0.0% | 8.9% |
|  | Rad | 12544 | 2.4% | 4.6% | 0.5% | 4.3% | 1.1% | 2.8% | 0.1% | 0.0% | 92.5% | 76.9% | 1.8% | 1.5% | 20.3% | 10.8% | 1.1% | 2.0% | 0.0% | 8.2% |
|  | UC | 3806 | 0.7% | 10.7% | 4.5% | 7.5% | 3.4% | 5.4% | 0.2% | 0.4% | 61.1% | 24.6% | 28.0% | 38.8% | 3.2% | 16.8% | 15.2% | 1.6% | 0.0% | 0.9% |
|  | All | 33343 | 6.4% | 8.5% | 2.8% | 6.4% | 2.8% | 4.4% | 0.2% | 0.2% | 78.1% | 48.2% | 22.3% | 26.4% | 11.4% | 23.5% | 16.2% | 2.2% | 0.0% | 7.7% |

PCP=Primary Care Provider, PA=Physician Assistant, DO=Doctor of Osteopathy, DC=Doctor of Chiropractic, PT=Physical Therapist, LAc=Licensed Acupuncturist, OS=Orthpedic Surgeon, PMR=Physical Medicine & Rehabilitation, PM=Pain Management, NS=Neurosurgeon, Neuro=Neurologist, Rheum=Rheumatologist, MD Oth=Other MD specialty, EM=Emergency Medicine, Rad=Radiologist, UC=Urgent Care
Cells with red text denote that the effect of provider type on service usage was found not to be significantly different from that of PCP-reference (Fisher's Exact p > 0.001)
Cells with black text denote that the effect of provider type on service usage was found to be significantly different from that of PCP-reference (Fisher's Exact p < 0.001)

**Table 2b.**
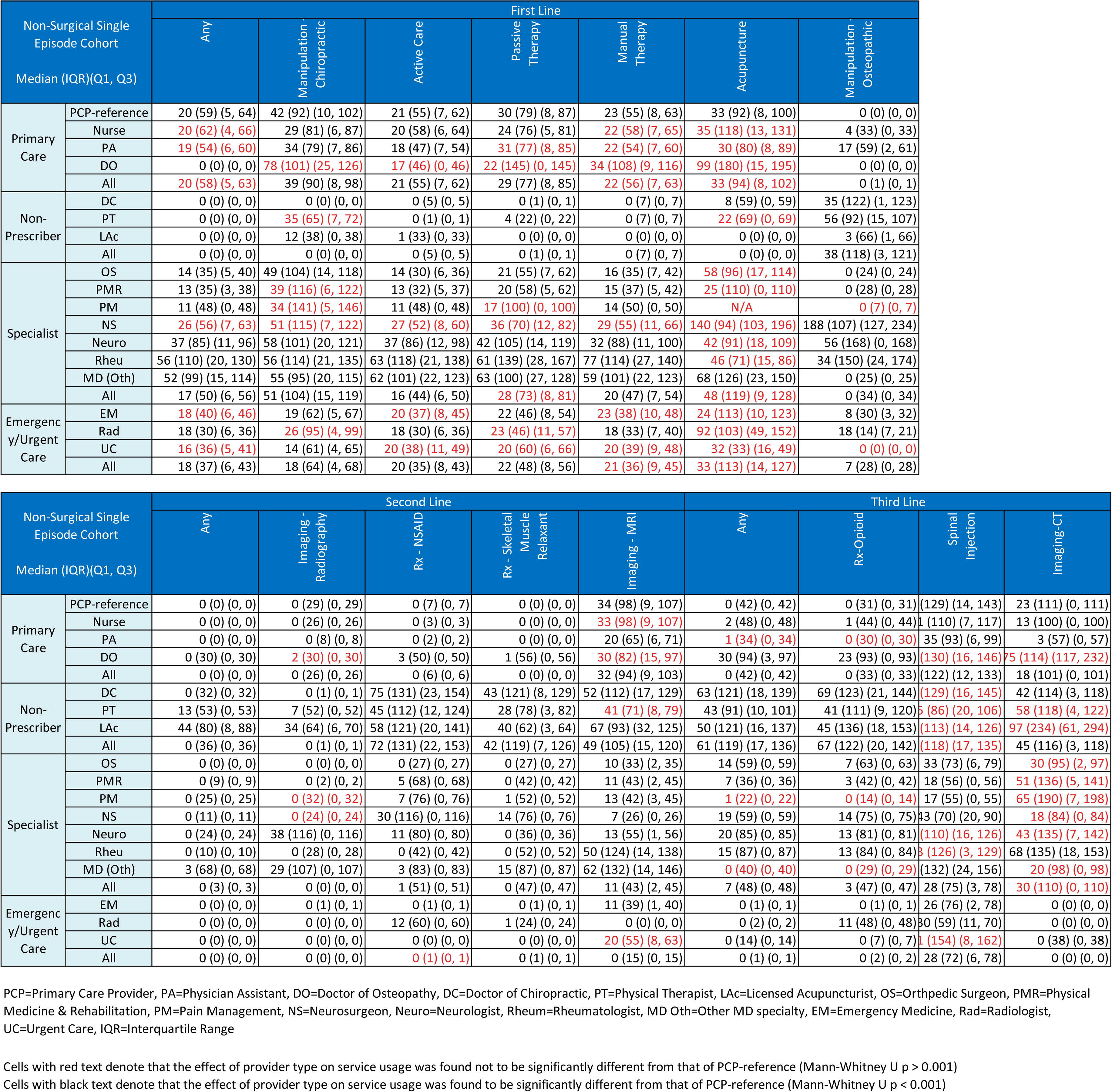
Low back pain # of days into episode when service initially provided by type of initial contact health care provider (HCP)

Supplement 3 presents the percent of episodes including each type of service for the overall non-surgical sample and Supplement 4 presents the results for the overall pooled sample. Supplement – 3a and 4a contain RRs for the non-surgical and pooled samples. Timing data is presented for the overall non-surgical (Supplement 5) and pooled (Supplement 6) samples. Due to the volume of data associated with replicating Tables 2a and 2b, this information is not separately reported for each episode sequence cohort. Among episode sequence cohorts there was a high degree of homogeneity in the distribution of initial contact HCP. Additionally, for each type of initial contact HCP there was also a high degree of homogeneity among episode sequence cohorts in the rate of service use (Figure 5).

**Figure 5.**
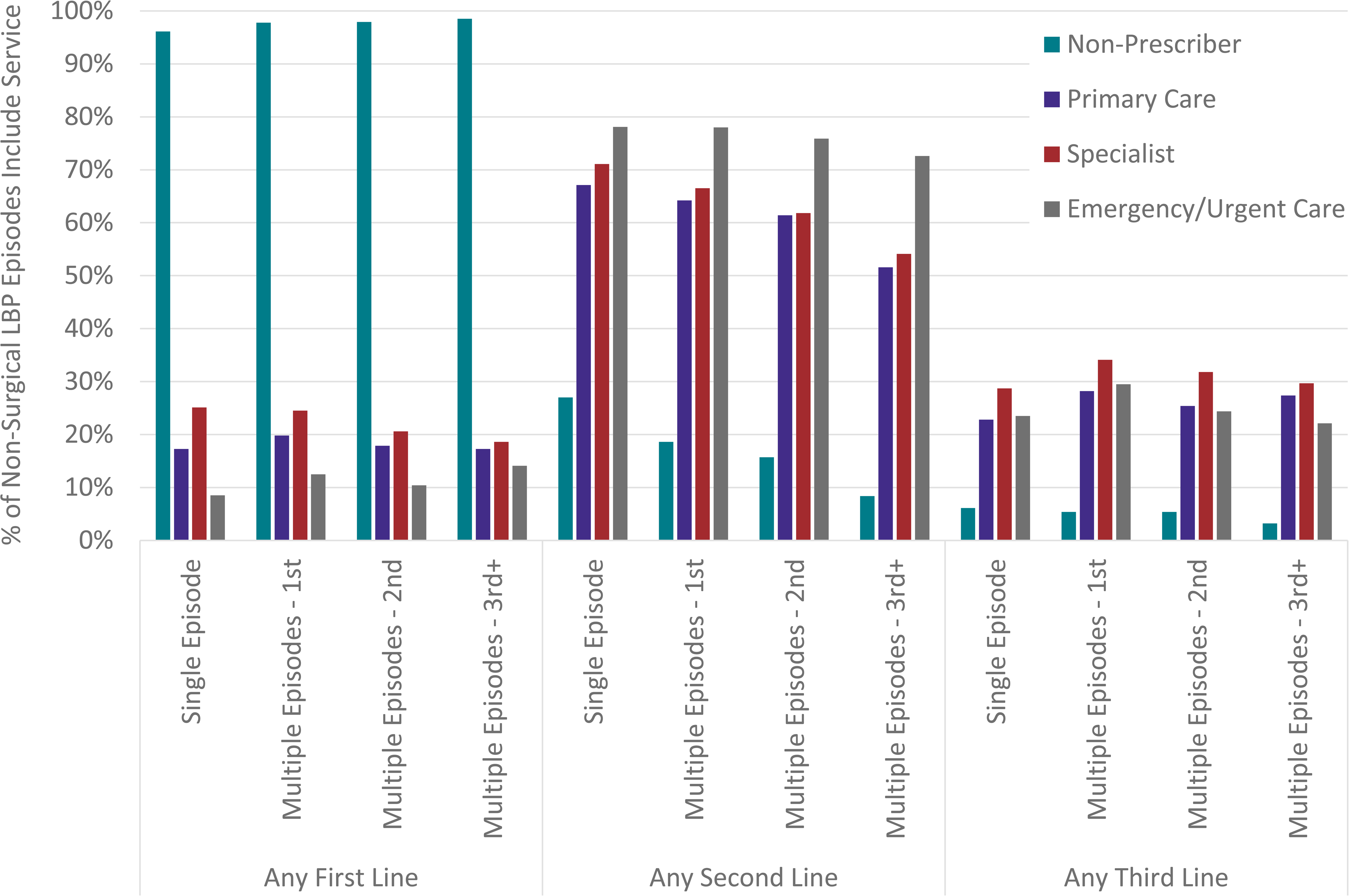
Percent of low back pain episodes including any first-, second- or third-line service by episode sequence cohort and type of initial contact health care provider

For the non-surgical sample, Table 2c presents total episode cost by episode sequence cohort and initial contact HCP. Supplement 7 presents this same data for the pooled sample. A mixed effects model with adjustment for covariates conducted on both pooled and non-surgical samples revealed virtually all HCPs were associated with significantly different total episode cost, opioid use and NSAID use than the PCP reference group (Table 3). As one example, for the adjusted non-surgical sample DCs had significantly lower total episode cost, with most specialist HCPs having significantly higher total episode cost compared to PCPs (Figure 6).

**Table 2c.** Low back pain total episode cost by episode sequence cohort

| Table 2c - Low back pain total episode cost by episode sequence cohort |  |  |  |  |  |  |
| --- | --- | --- | --- | --- | --- | --- |
| Median (IQR) (Q1, Q3) |  | Non-Surgical Episodes |  |  |  |  |
|  |  | Total | Single Episode | Multiple Episodes - First | Multiple Episodes - Second | Multiple Episodes - Third+ |
| Primary Care | PCP-reference | \$141 (353) (49, 401) | \$144 (352) (55, 407) | \$146 (380) (48, 428) | \$125 (354) (29, 383) | \$93 (212) (23, 235) |
| | Nurse | \$144 (419) (34, 453) | \$148 (415) (40, 455) | \$150 (457) (33, 490) | \$121 (416) (20, 436) | \$93 (237) (21, 258) |
| | PA | \$188 (553) (56, 609) | \$194 (541) (65, 606) | \$195 (637) (53, 690) | \$164 (551) (27, 578) | \$108 (314) (18, 333) |
| | DO | \$221 (518) (91, 608) | \$231 (501) (100, 601) | \$213 (600) (87, 687) | \$195 (554) (92, 646) | \$114 (162) (58, 220) |
| | All | \$145 (379) (47, 426) | \$149 (378) (54, 432) | \$149 (410) (47, 457) | \$128 (377) (27, 405) | \$94 (220) (23, 243) |
| Non-Prescriber | DC | \$165 (335) (65, 400) | \$185 (370) (80, 450) | \$160 (293) (67, 360) | \$135 (270) (60, 330) | \$90 (145) (50, 195) |
| | PT | \$718 (1164) (339, 1504) | \$694 (1119) (330, 1449) | \$906 (1634) (412, 2047) | \$719 (1128) (336, 1464) | \$560 (710) (338, 1048) |
| | LAc | \$363 (627) (156, 784) | \$353 (604) (146, 750) | \$381 (639) (183, 822) | \$400 (728) (185, 913) | \$365 (551) (156, 707) |
| | All | \$175 (352) (68, 420) | \$198 (401) (88, 489) | \$165 (316) (69, 385) | \$140 (290) (60, 350) | \$95 (148) (52, 200) |
| Specialist | OS | \$361 (947) (154, 1101) | \$360 (924) (159, 1083) | \$425 (1069) (160, 1228) | \$331 (1018) (123, 1141) | \$235 (747) (105, 853) |
| | PMR | \$681 (1619) (231, 1850) | \$689 (1603) (240, 1843) | \$708 (1699) (231, 1929) | \$652 (1651) (210, 1861) | \$365 (1238) (128, 1366) |
| | PM | \$587 (1568) (178, 1747) | \$643 (1700) (188, 1888) | \$541 (1319) (178, 1498) | \$508 (1363) (158, 1521) | \$271 (618) (143, 761) |
| | NS | \$816 (1867) (254, 2121) | \$861 (1888) (261, 2149) | \$820 (1937) (267, 2204) | \$690 (1596) (225, 1821) | \$504 (1526) (151, 1677) |
| | Neuro | \$468 (1259) (140, 1399) | \$535 (1369) (166, 1535) | \$444 (1199) (144, 1343) | \$332 (978) (101, 1079) | \$140 (508) (30, 537) |
| | Rheu | \$309 (831) (100, 931) | \$322 (885) (100, 985) | \$339 (982) (117, 1100) | \$271 (647) (93, 740) | \$247 (517) (74, 590) |
| | MD (Oth) | \$130 (501) (20, 521) | \$144 (570) (22, 593) | \$146 (515) (22, 537) | \$90 (322) (15, 337) | \$75 (225) (16, 241) |
| | All | \$389 (1125) (138, 1263) | \$404 (1134) (147, 1281) | \$426 (1205) (144, 1349) | \$324 (1061) (104, 1165) | \$221 (698) (74, 772) |
| Emergency/<br>Urgent Care | EM | \$550 (1362) (166, 1528) | \$564 (1359) (177, 1535) | \$586 (1407) (155, 1562) | \$415 (1384) (68, 1453) | \$319 (1213) (79, 1292) |
| | Rad | \$184 (536) (55, 590) | \$177 (507) (54, 561) | \$246 (812) (64, 876) | \$194 (655) (52, 708) | \$164 (631) (63, 695) |
| | UC | \$189 (253) (104, 356) | \$188 (241) (109, 349) | \$215 (273) (115, 388) | \$174 (277) (58, 335) | \$205 (358) (33, 391) |
| | All | \$303 (946) (93, 1038) | \$305 (936) (96, 1031) | \$355 (1079) (103, 1183) | \$253 (903) (56, 959) | \$242 (814) (64, 878) |

PCP=Primary Care Provider, PA=Physician Assistant, DO=Doctor of Osteopathy, DC=Doctor of Chiropractic, PT=Physical Therapist, LAc=Licensed Acupuncturist, OS=Orthpedic Surgeon, PMR=Physical Medicine & Rehabilitation, PM=Pain Management, NS=Neurosurgeon, Neuro=Neurologist, Rheum=Rheumatologist, MD Oth=Other MD specialty, EM=Emergency Medicine, Rad=Radiologist, UC=Urgent Care, IQR=Interquartile Range
Cells with red text denote that the effect of provider type on service usage was found not to be significantly different from that of PCP-reference (Mann-Whitney U p > 0.001) Cells with black text denote that the effect of provider type on service usage was found to be significantly different from that of PCP-reference (Mann-Whitney U p < 0.001)

**Figure 6.**
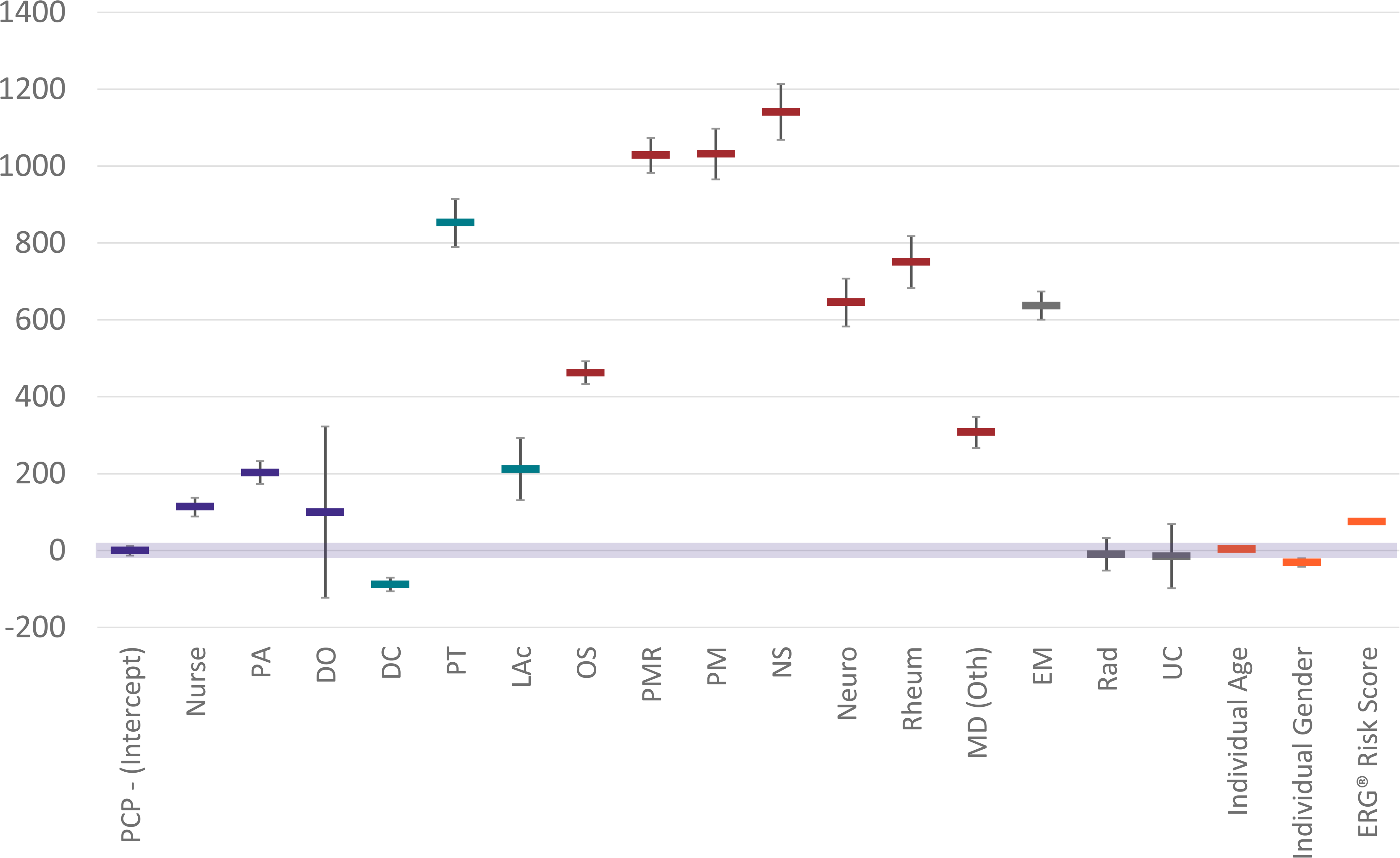
Low back pain non-surgical total episode cost regression estimates and 95% confidence intervals by type of initial contact HCP (0 Intercept = PCP reference group). PCP=Primary Care Provider; PA=Physician Assistant; DO=Doctor of Osteopathy; DC=Doctor of Chiropractic; PT=Physical Therapist; LAc=Licensed Acupuncturist; OS=Orthopedic Surgeon; PMR=Physical Medicine and Rehabilitation; PM=Pain Management, NS=Neurosurgeon; Neuro=Neurologist; Rheum=Rheumatologist; MD (Oth)=Other medical physician specialties, EM=Emergency Medicine; Rad=Radiologist; UC=Urgent Care

**Table 3.** Low back pain mixed effects model

| Features | Total Episode Cost |  | % of Episodes With Opioid |  | % of Episodes With NSAID |  |
| --- | --- | --- | --- | --- | --- | --- |
|  | Pooled | Non-Surgical | Pooled | Non-Surgical | Pooled | Non-Surgical |
| PCP - (Intercept) | 501 (460, 543) | 404 (391, 416) | 0.191 (0.189, 0.193) | 0.184 (0.182, 0.186) | 0.337 (0.335, 0.340) | 0.335 (0.332, 0.337) |
| Nurse | 165 (85, 245) | 114 (89, 138) | -0.022 (-0.026, -0.018) | -0.023 (-0.027, -0.019) | 0.024 (0.019, 0.028) | 0.024 (0.020, 0.028) |
| PA | 398 (302, 494) | 203 (173, 233) | -0.004 (-0.009, 0.001) | -0.009 (-0.014, -0.004) | 0.011 (0.005, 0.016) | 0.012 (0.006, 0.017) |
| DO | -377 (-1065, 312) | 100 (-122, 323) | -0.128 (-0.166, -0.090) | -0.119 (-0.157, -0.081) | -0.220 (-0.263, -0.178) | -0.217 (-0.260, -0.174) |
| DC | -322 (-377, -267) | -88 (-106, -70) | -0.163 (-0.166, -0.160) | -0.157 (-0.161, -0.154) | -0.275 (-0.279, -0.272) | -0.274 (-0.278, -0.271) |
| PT | 1359 (1155, 1563) | 853 (790, 915) | -0.116 (-0.126, -0.106) | -0.127 (-0.137, -0.118) | -0.200 (-0.211, -0.189) | -0.205 (-0.216, -0.194) |
| LAc | -45 (-311, 220) | 212 (131, 293) | -0.169 (-0.182, -0.156) | -0.164 (-0.177, -0.151) | -0.283 (-0.298, -0.268) | -0.280 (-0.295, -0.265) |
| OS | 2102 (2009, 2195) | 463 (433, 493) | -0.059 (-0.064, -0.054) | -0.080 (-0.085, -0.075) | -0.052 (-0.057, -0.046) | -0.056 (-0.061, -0.050) |
| PMR | 1885 (1748, 2022) | 1029 (983, 1074) | 0.002 (-0.006, 0.010) | -0.009 (-0.017, -0.001) | -0.107 (-0.115, -0.098) | -0.110 (-0.118, -0.101) |
| PM | 889 (690, 1089) | 1032 (966, 1098) | 0.059 (0.048, 0.069) | 0.076 (0.065, 0.087) | -0.193 (-0.204, -0.181) | -0.187 (-0.200, -0.175) |
| NS | 7664 (7457, 7870) | 1141 (1069, 1214) | 0.032 (0.021, 0.043) | -0.062 (-0.074, -0.051) | -0.152 (-0.164, -0.140) | -0.154 (-0.168, -0.141) |
| Neuro | 1214 (1013, 1415) | 646 (583, 708) | -0.061 (-0.071, -0.051) | -0.067 (-0.077, -0.058) | -0.097 (-0.108, -0.086) | -0.097 (-0.108, -0.085) |
| Rheum | 687 (471, 903) | 751 (683, 818) | -0.059 (-0.070, -0.048) | -0.063 (-0.074, -0.052) | -0.040 (-0.052, -0.028) | -0.039 (-0.052, -0.027) |
| MD (Oth) | 158 (21, 295) | 308 (267, 348) | 0.034 (0.027, 0.040) | 0.038 (0.032, 0.045) | -0.105 (-0.113, -0.098) | -0.104 (-0.111, -0.097) |
| EM | 535 (415, 655) | 637 (601, 674) | 0.080 (0.074, 0.086) | 0.080 (0.074, 0.085) | 0.030 (0.023, 0.037) | 0.029 (0.023, 0.036) |
| Rad | -644 (-784, -504) | -10 (-52, 33) | -0.226 (-0.232, -0.219) | -0.215 (-0.221, -0.208) | -0.328 (-0.336, -0.321) | -0.324 (-0.332, -0.317) |
| UC | -141 (-408, 125) | -15 (-98, 69) | -0.015 (-0.029, -0.001) | -0.014 (-0.028, -0.000) | -0.014 (-0.029, 0.002) | -0.013 (-0.029, 0.003) |
| Individual Age | 15 (13, 16) | 4 (4, 5) | 0.001 (0.001, 0.001) | 0.001 (0.001, 0.001) | 0.001 (0.000, 0.001) | 0.001 (0.000, 0.001) |
| Individual Gender | -258 (-295, -221) | -31 (-42, -20) | -0.013 (-0.014, -0.011) | -0.008 (-0.010, -0.007) | -0.009 (-0.011, -0.008) | -0.009 (-0.010, -0.007) |
| ERG® Risk Score | 322 (315, 329) | 75 (73, 78) | 0.017 (0.017, 0.017) | 0.015 (0.014, 0.015) | 0.004 (0.004, 0.004) | 0.003 (0.003, 0.004) |

PCP=Primary Care Provider, PA=Physician Assistant, DO=Doctor of Osteopathy, DC=Doctor of Chiropractic, PT=Physical Therapist, LAc=Licensed Acupuncturist, OS=Orthopedic Surgeon, PMR=Physical Medicine & Rehabilitation, PM=Pain Management, NS=Neurosurgeon, Neuro=Neurologist, Rheum=Rheumatologist, MD Oth=Other MD specialty, EM=Emergency Medicine, Rad=Radiologist, UC=Urgent Care
Pooled = Combined surgical and non-surgical episodes
Values in red were not statistically significant ( $p > 0.05$ ) compared to intercept
Values in black were statistically significant ( $p > 0.05$ ) compared to intercept

## Discussion

This retrospective cohort study provides a comprehensive analysis of the association between the type of initial contact HCP, service utilization, and total episode cost for the treatment of LBP. PCPs and DCs were the most common types of HCP initially contacted by individuals with LBP. The mix and timing of services received by an individual with LBP has a strong association with the type of HCP initially contacted. Non-prescribing HCPs emphasize guideline recommended first-line services, with DCs having the lowest total episode cost of any HCP for both pooled and non-surgical samples. Specialist HCPs were associated with frequent use of low-value services, and high total episode cost. These findings were consistent for individuals experiencing single or multiple episodes during the study period.

As an observational study of associations, with no attempt to generate causal inferences, it remains important to reinforce limitations to consider. The cohort had continuous highly uniform commercial insurance coverage and the processing of administrative claims data resulting in the *ETG*^®^ and *ERG*^®^ outputs benefited from extensive quality and actuarial control measures applied to the source administrative claims database. Nonetheless, data errors, variability in benefit plan design and enrollee cost-sharing responsibility, and missing information were potential sources of confounding or bias. Although the commercial insurer HCP database is under continual validation through credentialing and network demographic data update processes, it may have included errors or missing information. Summarizing total episode cost has several potential limitations associated with insurance coverage, nature of network participation, and alternative reimbursement models. The data set did not describe a U.S representative sample. Nonetheless, individuals and episodes from all 50 states and most US territories were included.

There are potential limitations associated with using episode of care as the unit of analysis. As one example, for the 19% of individuals with multiple episodes during the study period the episode of care unit of analysis does not provide an aggregated view of all data during the study period. The episode sequence cohort analyses indicate the risk of severity confounding is low.

The long pre- and post-episode clean periods resulted in each episode being a distinct event. There is a high degree of homogeneity in both initial contact HCP and subsequent care delivery whether LBP is a single episode event, or a series of sequential episodes. While associated with potential limitations, using episode of care unit of measurement has potential translational benefits in supporting the transition from fee for service to value-based episodic bundled payment arrangements.

Another important limitation was the risk of selection bias due to the limited ability to control for individual preference of type of initial contact HCP, individual expectations or requests for specific health care services, and potentially meaningful differences in clinical complexity of individuals seeking treatment. This is relevant when comparing episode attributes associated with the type of initial contact HCP. We attempted to address this limitation by narrowing the study population with several exclusions, performed a separate analysis for a non-surgical sample, for both pooled and non-surgical samples performed a separate analysis for each episode sequence cohort, controlled for comorbidities using a *ERG^®^* score, and in adjusted mixed effects models included individual demographic variables and a random effect to address variation in decision-making among individual HCPs of the same type.

A translational limitation of this study is that enrollee, geographic, and environmental factors associated with social disadvantage, race/ethnicity and HCP availability are important for understanding and addressing observed variability in the treatment of LBP. As an example, in our data the availability of non-prescribing HCPs generally has an inverse association with both the ADI and percent non-White population residing in a ZIP code. ZIP codes with lower availability of non-prescribing HCPs may be associated with higher rates of prescription medication use, including opioids and prescription NSAIDs. These findings were beyond the scope of this paper and will be addressed in a subsequent paper.

Our analyses corroborated and expanded upon the findings of earlier published work demonstrating the importance of the timing of access to non-prescribing HCPs for LBP. A recent national study found utilization of a range of health care services, and cost varied significantly based on the first HCP seen by an individual with LBP. The results of the well-controlled study are nearly identical to the associations observed in our study [43]. When a DC or PT is either the initial contact HCP or is consulted early during an episode of LBP, individuals were more likely to receive guideline-recommended management [25,27,44,45]. Initial contact with a DC has been shown to be associated with lower cost than most HCPs, while early access to physical therapy has had mixed total episode cost outcomes [25,27,46,47]. Our study also substantiated the findings of a recent narrative review that suggested that access to non-pharmacologic providers is associated with improved guideline concordance for treatment of spinal pain [48]. Reordering the sequencing of services in the early management of LBP has been proposed to systematically align clinical practice with the evidence-based CPGs [26,46,47,49]. Potential barriers to accessing non-pharmacologic health care services include the lack of benefit coverage or limitations on number of covered visits to a non-prescribing HCP, referral requirements, wait times for treatment, availability of transportation, the need to secure time away from work to participate in multiple visits, and individual characteristics [50, 51]. The development of delivery system and individual engagement strategies that prioritize initial contact with HCPs trained to administer non-pharmacologic therapies is a fertile area for future research and health care modeling.

Our study suggests there may be health care system-level opportunities to improve the value of LBP treatment. This is consistent with previous studies identifying the structure of the health care system as a barrier to aligning clinical practice with evidence-based care [48]. Specifically, two decision points within an episode of LBP appear important for improving guideline concordance and cost of care. The first is helping individuals directly access an initial HCP more likely to provide guideline-concordant care. The second is making it easier for primary care and specialist HCPs, when initially consulted by an individual with LBP without serious underlying pathology, to refer for guideline-concordant first-line services, before providing second- or third-line services. Either scenario necessitates an effective and timely process for triaging and referring individuals for guideline-concordant care.

For LBP episodes initially contacting a primary care or specialist HCP, our study reveals a low proportion of episodes include timely incorporation of guideline-adherent non-pharmacologic therapies. This is consistent with previous studies that found the cost of non-pharmacologic therapies and the additional administrative burden associated with referring individuals to a non-prescribing HCP are perceived as barriers by primary care physicians [52, 53]. Future research may be helpful to identify additional potential barriers to timely referral from primary care and specialist HCPs for recommended first-line non-pharmacologic services. Potential referral barriers may include inability of individuals to schedule a timely appointment with non-prescribing HCPs, absence of electronic health record interoperability to enable efficient referrals, concerns about lack of or limited insurance coverage for non-pharmacologic therapies, and variability in guideline concordance among individual non-prescribing HCPs leaving primary care and specialist HCPs uncertain to whom an individual should be referred.

## Conclusions

The HCP initially contacted by an individual with LBP may profoundly impact the services provided and cost of care. The results of this observational study suggest that in cases of LBP without serious pathology, non-guideline-concordant low-value treatments are frequent and associated with early contact with primary care and specialist HCPs. LBP episodes initially contacting DCs, LAcs, PTs, or DOs providing spinal manipulation, are more likely to be associated with guideline-concordant care. DCs were the most cost-efficient HCP for the treatment of LBP. With demographic differences present among individuals with LBP initially contacting different types of HCP, and unmeasured confounders such as patient preference, clinical complexity, and local environmental factors it is important not to over interpret results. Systemic changes across the health care delivery system should be considered to increase the likelihood of individuals seeking and receiving guideline-concordant, high-value care for LBP. In the absence of red flags this may include increasing the proportion of LBP episodes initially contacting a non-prescribing HCP and increasing primary care and specialist referrals for non-pharmacological treatment before introducing second- and third-line services.

## Supporting information

Supplement 1 - Individual State

Supplement 2 - Episode Sequence Cohort

Supplement 3 - Non-surgical Percent

Supplement 3a - Non-surgical Risk Ratio

Supplement 4 - Pooled Percent

Supplement 4a - Pooled Risk Ratio

Supplement 5 - Non-surgical Timing

Supplement 6 - Pooled Timing

Supplement 7 - Pooled Cost

Supplement - Care Pathways

Supplement - Cohort

Supplement - STROBE Checklist

## Data Availability

All data produced in the present study are available upon reasonable request to the authors

## List of Abbreviations

LBP: Low back pain
US: United States
CPG: Clinical practice guideline
DC: Doctor of Chiropractic
PT: Physical Therapist
EM: Emergency Medicine
HCP: Health care provider
LAc: Licensed Acupuncturist
ADI: Area Deprivation Index
STROBE: Strengthening the Reporting of Observational Studies in Epidemiology
ETG*^®^*: Episode Treatment Group*^®^*
ERG*^®^*: Episode Risk Group*^®^*
ACP: American College of Physicians
DO: Doctor of Osteopathy
OTC: Over the counter
MICE: Multiple imputation for chained equations
SD: Standard deviation
IQR: Interquartile range
OR: Odds ratio
RR: Risk ratio
PA: Physician’s Assistant
PCP: Primary care provider
OS: Orthopedic Surgeon
NS: Neurosurgeon
PMR: Physical Medicine and Rehabilitation
PM: Pain Management
UC: Urgent Care
Neuro: Neurologist
Rheum: Rheumatologist
CMT: Chiropractic manipulative treatment
OMT: Osteopathic manipulative treatment

## Declarations

### Ethics approval and consent to participate

The UnitedHealth Group Office of Human Research Affairs Institutional Review Board

### Consent for publication

Not applicable

### Availability of data and materials

The data are proprietary and are not available for public use but, under certain conditions, may be made available to editors and their approved auditors under a data-use agreement to confirm the findings of the current study.

## Authors’ contributions

Study conception and design; **DE, TK, STS**. Data acquisition; **DE, MZ, YG, SS.** Data analysis and interpretation; **DE, MZ, TK, STS, PA, YG, SS, AF.** Draft or substantially revise manuscript; **DE, TK, PA, STS, AF.**

## Acknowledgements

Amy Okaya made important contributions to the final review and editing of the manuscript.

