## Supplement 1 - Individual State for "Low back pain care pathways and costs: association with the type of initial contact health care provider. A retrospective cohort study"

### Supplement 1 - Low back pain episodes by individual home address state

| State | Pooled | | Non Surgical | | % Surgical |
|---|---|---|---|---|---|
| | Episodes | % of Total | Episodes | % of Total | |
| Total | 756631 | 100.0% | 732917 | 100.0% | 3.1% |
| TX | 88977 | 11.8% | 85977 | 11.7% | 3.4% |
| FL | 70103 | 9.3% | 67910 | 9.3% | 3.1% |
| MO | 35489 | 4.7% | 34358 | 4.7% | 3.2% |
| OH | 35183 | 4.6% | 34027 | 4.6% | 3.3% |
| IL | 34674 | 4.6% | 33557 | 4.6% | 3.2% |
| WI | 33227 | 4.4% | 32157 | 4.4% | 3.2% |
| CA | 32646 | 4.3% | 31937 | 4.4% | 2.2% |
| MN | 30254 | 4.0% | 29267 | 4.0% | 3.3% |
| NC | 28930 | 3.8% | 28068 | 3.8% | 3.0% |
| CO | 25450 | 3.4% | 24524 | 3.3% | 3.6% |
| AZ | 23809 | 3.1% | 23150 | 3.2% | 2.8% |
| GA | 21937 | 2.9% | 21259 | 2.9% | 3.1% |
| MD | 20867 | 2.8% | 20217 | 2.8% | 3.1% |
| NY | 20803 | 2.7% | 20230 | 2.8% | 2.8% |
| VA | 19710 | 2.6% | 19207 | 2.6% | 2.6% |
| IN | 17963 | 2.4% | 17204 | 2.3% | 4.2% |
| LA | 15294 | 2.0% | 14840 | 2.0% | 3.0% |
| TN | 15166 | 2.0% | 14681 | 2.0% | 3.2% |
| IA | 14650 | 1.9% | 14234 | 1.9% | 2.8% |
| NE | 14356 | 1.9% | 13985 | 1.9% | 2.6% |
| PA | 12776 | 1.7% | 12339 | 1.7% | 3.4% |
| WA | 12206 | 1.6% | 11997 | 1.6% | 1.7% |
| NJ | 10284 | 1.4% | 10056 | 1.4% | 2.2% |
| OK | 9895 | 1.3% | 9507 | 1.3% | 3.9% |
| AR | 9582 | 1.3% | 9221 | 1.3% | 3.8% |
| MI | 8722 | 1.2% | 8464 | 1.2% | 3.0% |
| KS | 8606 | 1.1% | 8278 | 1.1% | 3.8% |
| MS | 8403 | 1.1% | 8104 | 1.1% | 3.6% |
| OR | 7957 | 1.1% | 7817 | 1.1% | 1.8% |
| KY | 7342 | 1.0% | 7056 | 1.0% | 3.9% |
| UT | 7025 | 0.9% | 6719 | 0.9% | 4.4% |
| MA | 6651 | 0.9% | 6443 | 0.9% | 3.1% |
| SC | 6361 | 0.8% | 6112 | 0.8% | 3.9% |
| CT | 6310 | 0.8% | 6108 | 0.8% | 3.2% |
| RI | 5184 | 0.7% | 5052 | 0.7% | 2.5% |
| AL | 3811 | 0.5% | 3627 | 0.5% | 4.8% |
| NV | 3739 | 0.5% | 3601 | 0.5% | 3.7% |
| NM | 2826 | 0.4% | 2762 | 0.4% | 2.3% |
| DC | 2279 | 0.3% | 2235 | 0.3% | 1.9% |
| ND | 1988 | 0.3% | 1935 | 0.3% | 2.7% |
| WV | 1645 | 0.2% | 1589 | 0.2% | 3.4% |
| NH | 1290 | 0.2% | 1242 | 0.2% | 3.7% |
| ID | 1150 | 0.2% | 1120 | 0.2% | 2.6% |
| ME | 978 | 0.1% | 952 | 0.1% | 2.7% |
| SD | 957 | 0.1% | 923 | 0.1% | 3.6% |
| WY | 824 | 0.1% | 796 | 0.1% | 3.4% |
| DE | 775 | 0.1% | 737 | 0.1% | 4.9% |
| VI | 586 | 0.1% | 576 | 0.1% | 1.7% |
| MT | 488 | 0.1% | 471 | 0.1% | 3.5% |
| VT | 143 | 0.0% | 137 | 0.0% | 4.2% |
| AK | 107 | 0.0% | 103 | 0.0% | 3.7% |
| HI | 92 | 0.0% | 89 | 0.0% | 3.3% |
| PR | 89 | 0.0% | 88 | 0.0% | 1.1% |
| Unknown | 6072 | 0.8% | 5872 | 0.8% | 3.3% |

Pooled = Combined surgical and non-surgical episodes
