## Supplement 2 - Episode Sequence Cohort for "Low back pain care pathways and costs: association with the type of initial contact health care provider. A retrospective cohort study"

| Supplement 2 - Low back pain episode sequence cohorts | | | | | | | | | | |
|---|---|---|---|---|---|---|---|---|---|---|
| Individuals | | | Episode Sequence Cohorts | | | | Time Intervals (days) - Median (Interquartile Range - Q1,Q3) (Minimum) | | | |
| | Individuals | % | | | Episodes | % | Episode Duration | Clean Period - Before Initial | Clean Period - Between Sequential | Clean Period - After Final |
| 1 Episode | 500987 | 81.2% | Single Episode | | 500987 | 66.2% | 25 (1, 138) (1) | 640 (425, 863) (91) | N/A | 411 (252, 649) (61) |
| 2 Episodes | 96325 | 15.6% | Multiple Episodes | First | 115779 | 15.3% | 41 (1, 152) (1) | 367 (200, 578) (91) | N/A | N/A |
| 3 Episodes | 16136 | 2.6% | | Second | 115779 | 15.3% | 45 (1, 177) (1) | N/A | 207 (121, 343) (61) | 384 (216, 512) (61) |
| 4+ Episodes | 3318 | 0.5% | | Third+ | 24086 | 3.2% | 21 (1, 98) (1) | N/A | 167 (108, 270) (61) | 237 (134, 377) (61) |
| Total | 616766 | 100.0% | Total | | 756631 | 100.0% | 29 (1, 146) (1) | 585 (346, 832) (91) | 196 (118, 326) (61) | 398 (236, 607) (61) |
