## Supplement 3 - Non-surgical Percent for "Low back pain care pathways and costs: association with the type of initial contact health care provider. A retrospective cohort study"

### Supplement 3 - Low back pain % of episodes including service by type of initial contact health care provider (HCP)

| All Non-Surgical Episodes | | Episodes | | First Line | | | | | | | Second Line | | | | | Third Line | | | | |
|---|---|---|---|---|---|---|---|---|---|---|---|---|---|---|---|---|---|---|---|---|
| | | Count | % | Any | Manipulation - Chiropractic | Active Care | Passive Therapy | Manual Therapy | Acupuncture | Manipulation - Osteopathic | Any | Imaging - Radiography | Rx - NSAID | Rx - Skeletal Muscle Relaxant | Imaging - MRI | Any | Rx-Opioid | Spinal Injection | Spinal Surgery | Imaging-CT |
| Total | | 732917 | 100.0% | 44.5% | 34.2% | 18.6% | 15.4% | 12.5% | 0.9% | 0.9% | 52.3% | 24.4% | 22.6% | 21.2% | 8.5% | 18.7% | 14.8% | 4.6% | 0.0% | 1.5% |
| Primary Care | PCP-reference | 262616 | 33.9% | 17.7% | 6.8% | 11.4% | 5.8% | 8.1% | 0.3% | 1.7% | 64.1% | 22.5% | 33.4% | 32.6% | 8.1% | 24.1% | 21.1% | 4.0% | 0.0% | 1.3% |
| | Nurse | 51639 | 6.7% | 16.9% | 8.1% | 10.9% | 5.9% | 7.0% | 0.2% | 0.4% | 69.0% | 23.5% | 35.6% | 38.9% | 7.5% | 23.0% | 19.0% | 5.1% | 0.0% | 1.6% |
| | PA | 33372 | 4.3% | 17.8% | 7.0% | 13.1% | 5.1% | 8.8% | 0.3% | 0.3% | 72.7% | 28.9% | 34.2% | 38.6% | 10.1% | 25.0% | 20.1% | 6.2% | 0.0% | 1.9% |
| | DO | 740 | 0.1% | 65.4% | 3.8% | 15.4% | 4.7% | 8.0% | 0.8% | 58.2% | 27.4% | 10.4% | 11.1% | 10.7% | 6.5% | 13.1% | 8.6% | 6.1% | 0.0% | 0.8% |
| | All | 348367 | 45.0% | 17.7% | 7.0% | 11.5% | 5.7% | 8.0% | 0.3% | 1.5% | 65.6% | 23.2% | 33.7% | 34.1% | 8.2% | 24.0% | 20.7% | 4.4% | 0.0% | 1.4% |
| Non-Prescriber | DC | 233178 | 30.1% | 96.8% | 93.1% | 28.8% | 34.5% | 17.7% | 0.6% | 0.2% | 21.9% | 15.3% | 5.6% | 3.9% | 2.3% | 5.4% | 4.2% | 1.2% | 0.0% | 0.3% |
| | PT | 6767 | 0.9% | 97.8% | 10.2% | 95.0% | 26.7% | 73.6% | 1.5% | 0.7% | 43.5% | 23.7% | 13.3% | 10.0% | 18.1% | 17.3% | 9.1% | 10.0% | 0.0% | 1.4% |
| | LAc | 4159 | 0.5% | 97.6% | 9.1% | 18.6% | 42.6% | 44.0% | 88.2% | 0.5% | 11.8% | 5.2% | 5.2% | 2.7% | 3.1% | 5.7% | 3.7% | 2.2% | 0.0% | 0.3% |
| | All | 244104 | 31.5% | 96.9% | 89.3% | 30.5% | 34.4% | 19.7% | 2.2% | 0.2% | 22.4% | 15.4% | 5.8% | 4.0% | 2.7% | 5.7% | 4.4% | 1.5% | 0.0% | 0.3% |
| Specialist | OS | 41087 | 5.3% | 27.4% | 5.5% | 23.8% | 8.2% | 16.7% | 0.3% | 0.6% | 82.0% | 61.9% | 27.0% | 14.1% | 24.8% | 23.4% | 13.8% | 11.6% | 0.0% | 2.2% |
| | PMR | 18168 | 2.3% | 29.9% | 5.3% | 26.0% | 8.7% | 18.8% | 0.9% | 1.4% | 65.4% | 34.8% | 23.6% | 16.8% | 24.5% | 38.5% | 20.1% | 23.9% | 0.0% | 1.6% |
| | PM | 6922 | 0.9% | 9.0% | 1.7% | 7.5% | 2.7% | 5.0% | 0.1% | 0.5% | 47.4% | 14.6% | 16.6% | 17.2% | 17.0% | 55.1% | 32.0% | 31.0% | 0.0% | 1.5% |
| | NS | 5390 | 0.7% | 22.3% | 4.9% | 19.2% | 7.2% | 13.4% | 0.2% | 0.1% | 74.0% | 39.1% | 19.4% | 16.3% | 41.0% | 30.7% | 17.1% | 14.3% | 0.0% | 7.5% |
| | Neuro | 6628 | 0.9% | 19.5% | 7.3% | 14.5% | 6.6% | 9.6% | 0.4% | 0.4% | 62.9% | 15.6% | 24.6% | 25.2% | 27.3% | 25.6% | 17.6% | 9.7% | 0.0% | 2.5% |
| | Rheu | 6223 | 0.8% | 15.2% | 7.3% | 10.1% | 5.0% | 7.1% | 0.4% | 0.2% | 66.9% | 35.5% | 29.9% | 17.8% | 12.9% | 24.2% | 17.9% | 8.4% | 0.0% | 1.5% |
| | MD (Oth) | 14574 | 1.9% | 21.7% | 13.2% | 11.3% | 7.3% | 7.9% | 0.7% | 1.3% | 46.4% | 18.6% | 23.8% | 16.3% | 8.7% | 30.5% | 26.7% | 4.8% | 0.0% | 2.0% |
| | All | 98992 | 12.8% | 24.1% | 6.5% | 19.5% | 7.4% | 13.7% | 0.5% | 0.8% | 68.6% | 41.3% | 24.8% | 16.2% | 22.2% | 30.0% | 18.8% | 14.0% | 0.0% | 2.3% |
| Emergency/Urgent Care | EM | 21006 | 2.7% | 11.8% | 4.8% | 8.2% | 4.2% | 5.7% | 0.2% | 0.3% | 71.5% | 31.6% | 36.7% | 41.1% | 7.2% | 35.2% | 28.4% | 2.7% | 0.0% | 8.7% |
| | Rad | 15708 | 2.0% | 4.8% | 0.6% | 4.4% | 1.2% | 2.8% | 0.1% | 0.0% | 91.6% | 74.3% | 2.1% | 1.6% | 22.1% | 11.1% | 1.3% | 2.4% | 0.0% | 7.9% |
| | UC | 4740 | 0.6% | 11.3% | 4.9% | 7.5% | 3.6% | 5.4% | 0.2% | 0.5% | 60.8% | 23.7% | 28.2% | 38.2% | 3.6% | 18.3% | 16.5% | 2.0% | 0.0% | 0.8% |
| | All | 41454 | 5.4% | 9.1% | 3.2% | 6.7% | 3.0% | 4.6% | 0.2% | 0.2% | 77.9% | 46.9% | 22.6% | 25.8% | 12.4% | 24.1% | 16.8% | 2.5% | 0.0% | 7.5% |

PCP=Primary Care Provider, PA=Physician Assistant, DO=Doctor of Osteopathy, DC=Doctor of Chiropractic, PT=Physical Therapist, LAc=Licensed Acupuncturist, OS=Orthpedic Surgeon, PMR=Physical Medicine & Rehabilitation, PM=Pain Management, NS=Neurosurgeon, Neuro=Neurologist, Rheum=Rheumatologist, MD Oth=Other MD specialty, EM=Emergency Medicine, Rad=Radiologist, UC=Urgent Care

Cells with red text denote that the effect of provider type on service usage was found not to be significantly different from that of PCP-reference (Fisher's Exact $p > 0.001$)
Cells with black text denote that the effect of provider type on service usage was found to be significantly different from that of PCP-reference (Fisher's Exact $p < 0.001$)
