## Supplement 3a - Non-surgical Risk Ratio for "Low back pain care pathways and costs: association with the type of initial contact health care provider. A retrospective cohort study"

### Supplement 3a - Risk ratio and 95% confidence interval for service use during non-surgical low back pain episodes by type of initial contact health care provider as compared to a primary care provider (PCP)

| All Non-Surgical Episodes | | First Line | | | | | | | Second Line | | | | | Third Line | | | | |
|---|---|---|---|---|---|---|---|---|---|---|---|---|---|---|---|---|---|---|
| | | Any | Manipulation - Chiropractic | Active Care | Passive Therapy | Manual Therapy | Acupuncture | Manipulation - Osteopathic | Any | Imaging - Radiography | Rx - NSAID | Rx - Skeletal Muscle Relaxant | Imaging - MRI | Any | Rx-Opioid | Spinal Injection | Spinal Surgery | Imaging-CT |
| Primary Care | PCP | reference | | | | | | | | | | | | | | | | |
| | Nurse | 0.95 (0.93, 0.97) | 1.19 (1.15, 1.23) | 0.95 (0.93, 0.98) | 1.03 (0.99, 1.06) | 0.87 (0.84, 0.90) | 0.57 (0.45, 0.71) | 0.26 (0.23, 0.30) | 1.08 (1.07, 1.08) | 1.04 (1.03, 1.06) | 1.07 (1.05, 1.08) | 1.19 (1.18, 1.21) | 0.93 (0.90, 0.96) | 0.96 (0.94, 0.97) | 0.90 (0.89, 0.92) | 1.26 (1.21, 1.32) | N/A | 1.27 (1.18, 1.37) |
| | PA | 1.01 (0.98, 1.03) | 1.03 (0.98, 1.07) | 1.14 (1.11, 1.18) | 0.89 (0.85, 0.93) | 1.08 (1.04, 1.12) | 1.08 (0.88, 1.34) | 0.18 (0.15, 0.22) | 1.13 (1.13, 1.14) | 1.29 (1.26, 1.31) | 1.02 (1.01, 1.04) | 1.18 (1.17, 1.20) | 1.25 (1.20, 1.29) | 1.04 (1.02, 1.06) | 0.96 (0.93, 0.98) | 1.55 (1.48, 1.62) | N/A | 1.54 (1.42, 1.68) |
| | DO | 3.69 (3.50, 3.89) | 0.56 (0.39, 0.80) | 1.35 (1.14, 1.60) | 0.82 (0.59, 1.13) | 0.98 (0.77, 1.26) | 2.93 (1.32, 6.53) | 34.72 (32.44, | 0.43 (0.38, 0.48) | 0.46 (0.37, 0.57) | 0.33 (0.27, 0.41) | 0.33 (0.27, 0.40) | 0.80 (0.61, 1.05) | 0.54 (0.45, 0.66) | 0.41 (0.32, 0.52) | 1.52 (1.14, 2.02) | N/A | 0.65 (0.29, 1.43) |
| Non-Prescriber | DC | 5.46 (5.41, 5.50) | 13.65 (13.46, 13.85) | 2.52 (2.49, 2.55) | 5.97 (5.87, 6.07) | 2.18 (2.15, 2.22) | 2.32 (2.12, 2.53) | 0.09 (0.08, 0.10) | 0.34 (0.34, 0.35) | 0.68 (0.67, 0.69) | 0.17 (0.16, 0.17) | 0.12 (0.12, 0.12) | 0.28 (0.27, 0.29) | 0.22 (0.22, 0.23) | 0.20 (0.20, 0.20) | 0.30 (0.29, 0.32) | N/A | 0.25 (0.23, 0.27) |
| | PT | 5.52 (5.47, 5.56) | 1.49 (1.39, 1.61) | 8.31 (8.21, 8.41) | 4.61 (4.42, 4.81) | 9.09 (8.92, 9.26) | 5.29 (4.30, 6.52) | 0.39 (0.29, 0.52) | 0.68 (0.66, 0.70) | 1.05 (1.01, 1.10) | 0.40 (0.37, 0.42) | 0.31 (0.28, 0.33) | 2.23 (2.12, 2.35) | 0.72 (0.68, 0.76) | 0.43 (0.40, 0.47) | 2.50 (2.32, 2.69) | N/A | 1.13 (0.92, 1.38) |
| | LAc | 5.50 (5.45, 5.56) | 1.34 (1.21, 1.47) | 1.63 (1.53, 1.74) | 7.36 (7.08, 7.65) | 5.44 (5.24, 5.64) | 319.11 (296.50, 343.45) | 0.27 (0.17, 0.43) | 0.18 (0.17, 0.20) | 0.23 (0.20, 0.26) | 0.16 (0.14, 0.18) | 0.08 (0.07, 0.10) | 0.38 (0.32, 0.45) | 0.23 (0.21, 0.27) | 0.18 (0.15, 0.21) | 0.54 (0.44, 0.66) | N/A | 0.21 (0.12, 0.38) |
| Specialist | OS | 1.54 (1.52, 1.57) | 0.81 (0.77, 0.84) | 2.08 (2.04, 2.12) | 1.42 (1.37, 1.47) | 2.06 (2.01, 2.12) | 0.97 (0.79, 1.18) | 0.34 (0.30, 0.39) | 1.28 (1.27, 1.29) | 2.76 (2.73, 2.78) | 0.81 (0.80, 0.82) | 0.43 (0.42, 0.44) | 3.06 (3.00, 3.13) | 0.97 (0.95, 0.99) | 0.66 (0.64, 0.67) | 2.89 (2.80, 2.99) | N/A | 1.72 (1.60, 1.85) |
| | PMR | 1.68 (1.64, 1.72) | 0.78 (0.73, 0.83) | 2.27 (2.21, 2.34) | 1.50 (1.43, 1.58) | 2.32 (2.24, 2.40) | 3.42 (2.90, 4.04) | 0.86 (0.76, 0.97) | 1.02 (1.01, 1.03) | 1.55 (1.52, 1.58) | 0.71 (0.69, 0.73) | 0.51 (0.50, 0.53) | 3.02 (2.94, 3.11) | 1.60 (1.57, 1.63) | 0.96 (0.93, 0.98) | 5.96 (5.77, 6.15) | N/A | 1.24 (1.10, 1.40) |
| | PM | 0.51 (0.47, 0.55) | 0.24 (0.20, 0.29) | 0.65 (0.60, 0.71) | 0.47 (0.41, 0.55) | 0.62 (0.55, 0.68) | 0.26 (0.11, 0.63) | 0.31 (0.22, 0.43) | 0.74 (0.72, 0.76) | 0.65 (0.61, 0.69) | 0.50 (0.47, 0.52) | 0.53 (0.50, 0.56) | 2.10 (1.99, 2.21) | 2.29 (2.24, 2.34) | 1.52 (1.46, 1.57) | 7.75 (7.45, 8.07) | N/A | 1.16 (0.95, 1.41) |
| | Neuro | 1.26 (1.19, 1.32) | 0.71 (0.63, 0.80) | 1.68 (1.59, 1.78) | 1.25 (1.13, 1.37) | 1.66 (1.55, 1.77) | 0.67 (0.36, 1.25) | 0.08 (0.04, 0.16) | 1.15 (1.14, 1.17) | 1.74 (1.68, 1.80) | 0.58 (0.55, 0.61) | 0.50 (0.47, 0.53) | 5.05 (4.88, 5.23) | 1.28 (1.23, 1.33) | 0.81 (0.76, 0.86) | 3.58 (3.34, 3.83) | N/A | 5.94 (5.37, 6.56) |
| | Neurologist | 1.10 (1.05, 1.16) | 1.07 (0.98, 1.17) | 1.27 (1.20, 1.35) | 1.14 (1.04, 1.25) | 1.18 (1.10, 1.28) | 1.58 (1.09, 2.29) | 0.22 (0.14, 0.32) | 0.98 (0.96, 1.00) | 0.69 (0.66, 0.73) | 0.74 (0.71, 0.77) | 0.77 (0.74, 0.80) | 3.37 (3.23, 3.51) | 1.06 (1.02, 1.11) | 0.83 (0.79, 0.88) | 2.43 (2.25, 2.62) | N/A | 1.98 (1.70, 2.31) |
| | Rheu | 0.86 (0.81, 0.91) | 1.08 (0.98, 1.18) | 0.89 (0.82, 0.96) | 0.86 (0.77, 0.96) | 0.88 (0.80, 0.96) | 1.51 (1.02, 2.23) | 0.13 (0.08, 0.23) | 1.04 (1.03, 1.06) | 1.58 (1.52, 1.63) | 0.90 (0.86, 0.93) | 0.55 (0.52, 0.58) | 1.59 (1.49, 1.69) | 1.00 (0.96, 1.05) | 0.85 (0.81, 0.90) | 2.09 (1.92, 2.27) | N/A | 1.19 (0.97, 1.46) |
| | MD (Oth) | 1.23 (1.19, 1.26) | 1.93 (1.85, 2.02) | 0.98 (0.94, 1.03) | 1.26 (1.18, 1.33) | 0.97 (0.92, 1.03) | 2.43 (1.97, 3.00) | 0.76 (0.66, 0.88) | 0.72 (0.71, 0.74) | 0.83 (0.80, 0.86) | 0.71 (0.69, 0.73) | 0.50 (0.48, 0.52) | 1.07 (1.01, 1.13) | 1.27 (1.24, 1.30) | 1.27 (1.23, 1.30) | 1.20 (1.11, 1.29) | N/A | 1.61 (1.43, 1.81) |
| Emergency/ Urgent Care | EM | 0.67 (0.64, 0.69) | 0.71 (0.66, 0.75) | 0.72 (0.69, 0.75) | 0.72 (0.68, 0.77) | 0.70 (0.66, 0.74) | 0.76 (0.56, 1.03) | 0.16 (0.12, 0.21) | 1.11 (1.10, 1.13) | 1.41 (1.38, 1.44) | 1.10 (1.08, 1.12) | 1.26 (1.24, 1.28) | 0.89 (0.85, 0.94) | 1.46 (1.43, 1.49) | 1.35 (1.32, 1.38) | 0.68 (0.62, 0.74) | N/A | 6.93 (6.56, 7.33) |
| | Rad | 0.27 (0.25, 0.29) | 0.09 (0.07, 0.11) | 0.38 (0.36, 0.41) | 0.21 (0.18, 0.24) | 0.35 (0.32, 0.38) | 0.30 (0.17, 0.52) | 0.03 (0.01, 0.06) | 1.43 (1.42, 1.44) | 3.31 (3.27, 3.35) | 0.06 (0.06, 0.07) | 0.05 (0.04, 0.06) | 2.72 (2.64, 2.81) | 0.46 (0.44, 0.48) | 0.06 (0.05, 0.07) | 0.60 (0.54, 0.66) | N/A | 6.33 (5.94, 6.74) |
| | UC | 0.64 (0.59, 0.69) | 0.71 (0.63, 0.81) | 0.65 (0.59, 0.72) | 0.63 (0.54, 0.73) | 0.67 (0.59, 0.75) | 0.84 (0.46, 1.52) | 0.31 (0.21, 0.47) | 0.95 (0.93, 0.97) | 1.05 (1.00, 1.11) | 0.85 (0.81, 0.89) | 1.17 (1.13, 1.21) | 0.44 (0.38, 0.51) | 0.76 (0.72, 0.81) | 0.78 (0.73, 0.83) | 0.51 (0.41, 0.62) | N/A | 0.67 (0.49, 0.92) |

PCP=Primary Care Provider, PA=Physician Assistant, DO=Doctor of Osteopathy, DC=Doctor of Chiropractic, PT=Physical Therapist, LAc=Licensed Acupuncturist, OS=Orthpedic Surgeon, PMR=Physical Medicine & Rehabilitation, PM=Pain Management, NS=Neurosurgeon, Neuro=Neurologist, Rheum=Rheumatologist, MD Oth=Other MD specialty, EM=Emergency Medicine, Rad=Radiologist, UC=Urgent Care

Cells in red are not different than the PCP reference (p=.05)
