## Supplement 4 - Pooled Percent for "Low back pain care pathways and costs: association with the type of initial contact health care provider. A retrospective cohort study"

### Supplement 4 - Low back pain % of episodes including service by type of initial contact health care provider (HCP)

| All Combined Surgical and Non-Surgical (Pooled) Episodes | | Episodes | | First Line | | | | | | | Second Line | | | | | Third Line | | | | |
|---|---|---|---|---|---|---|---|---|---|---|---|---|---|---|---|---|---|---|---|---|
| | | Count | % | Any | Manipulation - Chiropractic | Active Care | Passive Therapy | Manual Therapy | Acupuncture | Manipulation - Osteopathic | Any | Imaging - Radiography | Rx - NSAID | Rx - Skeletal Muscle Relaxant | Imaging - MRI | Any | Rx-Opioid | Spinal Injection | Spinal Surgery | Imaging-CT |
| Total | | 756631 | 100.0% | 44.5% | 33.5% | 19.3% | 15.4% | 12.9% | 0.9% | 0.9% | 53.5% | 25.6% | 23.1% | 21.9% | 10.3% | 21.3% | 16.2% | 6.7% | 3.1% | 1.8% |
| Primary Care | PCP-reference | 269925 | 33.8% | 18.5% | 6.9% | 12.3% | 6.1% | 8.6% | 0.3% | 1.7% | 64.9% | 23.6% | 33.8% | 33.1% | 9.8% | 26.1% | 22.3% | 5.9% | 2.7% | 1.5% |
| | Nurse | 53355 | 6.7% | 17.7% | 8.2% | 11.7% | 6.3% | 7.6% | 0.2% | 0.4% | 69.7% | 24.6% | 36.1% | 39.3% | 9.4% | 25.5% | 20.5% | 7.3% | 3.2% | 1.9% |
| | PA | 34781 | 4.4% | 18.9% | 7.1% | 14.1% | 5.5% | 9.4% | 0.3% | 0.3% | 73.5% | 30.3% | 34.5% | 39.0% | 12.4% | 28.1% | 22.0% | 8.9% | 4.1% | 2.3% |
| | DO | 747 | 0.1% | 65.3% | 3.7% | 15.4% | 4.7% | 7.9% | 0.9% | 58.1% | 28.0% | 11.0% | 11.2% | 10.8% | 6.8% | 13.9% | 9.0% | 6.6% | 0.9% | 0.9% |
| | All | 358808 | 44.9% | 18.5% | 7.1% | 12.4% | 6.1% | 8.5% | 0.3% | 1.5% | 66.4% | 24.4% | 34.2% | 34.6% | 10.0% | 26.2% | 22.0% | 6.4% | 2.9% | 1.7% |
| Non-Prescriber | DC | 234868 | 29.4% | 96.8% | 93.1% | 29.0% | 34.6% | 17.9% | 0.6% | 0.2% | 22.4% | 15.7% | 5.8% | 4.2% | 2.8% | 6.1% | 4.6% | 1.7% | 0.7% | 0.4% |
| | PT | 7143 | 0.9% | 97.8% | 10.2% | 95.0% | 27.4% | 73.7% | 1.5% | 0.7% | 45.9% | 25.6% | 14.4% | 11.1% | 20.6% | 21.7% | 11.5% | 13.8% | 5.3% | 2.0% |
| | LAc | 4193 | 0.5% | 97.6% | 9.3% | 18.9% | 42.7% | 44.2% | 88.2% | 0.5% | 12.4% | 5.6% | 5.3% | 3.0% | 3.6% | 6.4% | 4.2% | 2.8% | 0.8% | 0.3% |
| | All | 246204 | 30.8% | 96.9% | 89.2% | 30.8% | 34.6% | 19.9% | 2.2% | 0.2% | 23.0% | 15.8% | 6.1% | 4.3% | 3.3% | 6.5% | 4.8% | 2.1% | 0.9% | 0.4% |
| Specialist | OS | 45363 | 5.7% | 28.8% | 5.5% | 25.3% | 8.7% | 17.5% | 0.3% | 0.5% | 83.0% | 63.3% | 27.9% | 15.9% | 28.7% | 30.6% | 18.0% | 16.6% | 9.4% | 3.1% |
| | PMR | 19974 | 2.5% | 30.6% | 5.5% | 26.8% | 9.1% | 19.2% | 0.9% | 1.4% | 66.9% | 36.1% | 24.5% | 18.2% | 27.4% | 44.1% | 23.1% | 29.6% | 9.0% | 2.2% |
| | PM | 7958 | 1.0% | 9.4% | 1.7% | 7.9% | 2.9% | 5.2% | 0.1% | 0.5% | 48.1% | 15.4% | 16.9% | 17.6% | 18.7% | 60.9% | 32.5% | 39.5% | 13.0% | 1.8% |
| | NS | 7314 | 0.9% | 26.9% | 4.7% | 24.0% | 8.4% | 15.9% | 0.2% | 0.1% | 77.9% | 46.9% | 20.5% | 22.7% | 45.6% | 49.0% | 28.5% | 22.3% | 26.3% | 9.9% |
| | Neuro | 7115 | 0.9% | 20.6% | 7.3% | 15.7% | 7.1% | 10.3% | 0.4% | 0.4% | 64.5% | 17.7% | 25.1% | 26.2% | 29.6% | 30.7% | 20.1% | 14.0% | 6.8% | 3.2% |
| | Rheu | 6500 | 0.8% | 16.1% | 7.3% | 11.0% | 5.2% | 7.6% | 0.4% | 0.3% | 67.9% | 36.4% | 30.5% | 19.0% | 14.8% | 27.4% | 20.1% | 11.2% | 4.3% | 1.9% |
| | MD (Oth) | 15070 | 1.9% | 22.4% | 13.1% | 12.1% | 7.4% | 8.4% | 0.7% | 1.3% | 47.7% | 19.9% | 24.2% | 17.2% | 10.2% | 32.8% | 27.9% | 6.9% | 3.3% | 2.4% |
| | All | 109294 | 13.7% | 25.4% | 6.4% | 20.9% | 7.8% | 14.5% | 0.4% | 0.7% | 70.2% | 43.2% | 25.5% | 17.9% | 25.6% | 36.6% | 22.3% | 19.2% | 9.4% | 3.1% |
| Emergency/ Urgent Care | EM | 21545 | 2.7% | 12.8% | 5.0% | 9.1% | 4.5% | 6.2% | 0.2% | 0.3% | 72.1% | 32.7% | 37.1% | 41.5% | 9.1% | 36.8% | 29.6% | 4.3% | 2.5% | 9.0% |
| | Rad | 15971 | 2.0% | 5.1% | 0.6% | 4.6% | 1.3% | 2.9% | 0.1% | 0.0% | 91.5% | 73.7% | 2.2% | 1.8% | 22.9% | 12.5% | 1.7% | 3.7% | 1.6% | 8.0% |
| | UC | 4809 | 0.6% | 12.1% | 5.0% | 8.2% | 3.8% | 5.9% | 0.2% | 0.5% | 61.4% | 24.5% | 28.6% | 38.4% | 4.7% | 19.5% | 17.3% | 3.0% | 1.4% | 0.9% |
| | All | 42325 | 5.3% | 9.8% | 3.3% | 7.3% | 3.2% | 5.0% | 0.2% | 0.2% | 78.2% | 47.2% | 23.0% | 26.1% | 13.8% | 25.7% | 17.7% | 3.9% | 2.1% | 7.7% |

PCP=Primary Care Provider, PA=Physician Assistant, DO=Doctor of Osteopathy, DC=Doctor of Chiropractic, PT=Physical Therapist, LAc=Licensed Acupuncturist, OS=Orthpedic Surgeon, PMR=Physical Medicine & Rehabilitation, PM=Pain Management, NS=Neurosurgeon, Neuro=neurologist, Rheum=Rheumatologist, MD Oth=Other MD specialty, EM=Emergency Medicine, Rad=Radiologist, UC=Urgent Care

Cells with red text denote that the effect of provider type on service usage was found not to be significantly different from that of PCP-reference (Fisher's Exact $p > 0.001$)
Cells with black text denote that the effect of provider type on service usage was found to be significantly different from that of PCP-reference (Fisher's Exact $p < 0.001$)
