## Supplement 4a - Pooled Risk Ratio for "Low back pain care pathways and costs: association with the type of initial contact health care provider. A retrospective cohort study"

**Supplement 4a - Risk ratio and 95% confidence interval for service use during combined surgical and non-surgical (pooled) low back pain episodes by type of initial contact health care provider as compared to a primary care provider (PCP)**

| Combined Surgical and Non-surgical (Pooled) Episodes | | First Line | | | | | | | Second Line | | | | | Third Line | | | | |
|---|---|---|---|---|---|---|---|---|---|---|---|---|---|---|---|---|---|---|
| | | Any | Manipulation - Chiropractic | Active Care | Passive Therapy | Manual Therapy | Acupuncture | Manipulation - Osteopathic | Any | Imaging - Radiography | Rx - NSAID | Rx - Skeletal Muscle Relaxant | Imaging - MRI | Any | Rx-Opioid | Spinal Injection | Spinal Surgery | Imaging-CT |
| Primary Care | PCP | reference | | | | | | | | | | | | | | | | |
| | Nurse | 0.96 (0.94, 0.98) | 1.19 (1.15, 1.23) | 0.96 (0.93, 0.98) | 1.03 (0.99, 1.07) | 0.88 (0.85, 0.91) | 0.56 (0.45, 0.70) | 0.27 (0.24, 0.31) | 1.07 (1.07, 1.08) | 1.04 (1.03, 1.06) | 1.07 (1.05, 1.08) | 1.19 (1.17, 1.20) | 0.95 (0.93, 0.98) | 0.98 (0.96, 0.99) | 0.92 (0.90, 0.93) | 1.25 (1.21, 1.29) | 1.19 (1.13, 1.25) | 1.21 (1.13, 1.30) |
| | PA | 1.02 (1.00, 1.04) | 1.03 (0.99, 1.08) | 1.15 (1.12, 1.19) | 0.91 (0.87, 0.95) | 1.09 (1.05, 1.13) | 1.07 (0.87, 1.31) | 0.18 (0.15, 0.22) | 1.13 (1.12, 1.14) | 1.28 (1.26, 1.31) | 1.02 (1.01, 1.04) | 1.18 (1.16, 1.19) | 1.27 (1.23, 1.31) | 1.07 (1.06, 1.09) | 0.99 (0.96, 1.01) | 1.51 (1.46, 1.57) | 1.50 (1.41, 1.58) | 1.47 (1.37, 1.59) |
| | DO | 3.53 (3.35, 3.72) | 0.54 (0.38, 0.78) | 1.26 (1.06, 1.49) | 0.77 (0.56, 1.06) | 0.91 (0.72, 1.17) | 3.26 (1.55, 6.84) | 35.04 (32.76, 37.49) | 0.43 (0.38, 0.48) | 0.46 (0.38, 0.57) | 0.33 (0.27, 0.41) | 0.33 (0.27, 0.40) | 0.69 (0.53, 0.91) | 0.53 (0.45, 0.64) | 0.40 (0.32, 0.50) | 1.12 (0.85, 1.47) | 0.35 (0.17, 0.72) | 0.61 (0.29, 1.27) |
| Non-Prescriber | DC | 5.23 (5.19, 5.27) | 13.48 (13.30, 13.67) | 2.37 (2.34, 2.40) | 5.67 (5.58, 5.76) | 2.07 (2.04, 2.10) | 2.26 (2.07, 2.46) | 0.09 (0.08, 0.10) | 0.35 (0.34, 0.35) | 0.67 (0.66, 0.67) | 0.17 (0.17, 0.18) | 0.13 (0.12, 0.13) | 0.28 (0.28, 0.29) | 0.23 (0.23, 0.24) | 0.21 (0.20, 0.21) | 0.30 (0.29, 0.31) | 0.27 (0.25, 0.28) | 0.24 (0.22, 0.26) |
| | PT | 5.28 (5.24, 5.33) | 1.48 (1.38, 1.59) | 7.75 (7.66, 7.84) | 4.48 (4.30, 4.67) | 8.54 (8.39, 8.70) | 5.21 (4.26, 6.37) | 0.40 (0.30, 0.53) | 0.71 (0.69, 0.73) | 1.08 (1.04, 1.13) | 0.43 (0.40, 0.45) | 0.34 (0.31, 0.36) | 2.09 (2.00, 2.19) | 0.83 (0.79, 0.87) | 0.52 (0.48, 0.55) | 2.36 (2.22, 2.51) | 1.94 (1.76, 2.15) | 1.32 (1.12, 1.55) |
| | LAc | 5.27 (5.22, 5.32) | 1.35 (1.23, 1.49) | 1.54 (1.45, 1.64) | 7.00 (6.73, 7.27) | 5.12 (4.94, 5.31) | 306.69 (285.64, 329.30) | 0.27 (0.17, 0.43) | 0.19 (0.18, 0.21) | 0.24 (0.21, 0.27) | 0.16 (0.14, 0.18) | 0.09 (0.08, 0.11) | 0.36 (0.31, 0.43) | 0.25 (0.22, 0.28) | 0.19 (0.16, 0.22) | 0.48 (0.40, 0.57) | 0.30 (0.21, 0.42) | 0.19 (0.11, 0.33) |
| Specialist | OS | 1.56 (1.53, 1.58) | 0.80 (0.77, 0.84) | 2.07 (2.03, 2.11) | 1.42 (1.37, 1.47) | 2.02 (1.97, 2.07) | 0.94 (0.78, 1.14) | 0.33 (0.29, 0.38) | 1.28 (1.27, 1.29) | 2.68 (2.65, 2.71) | 0.83 (0.81, 0.84) | 0.48 (0.47, 0.49) | 2.93 (2.87, 2.98) | 1.17 (1.15, 1.19) | 0.81 (0.79, 0.82) | 2.83 (2.76, 2.90) | 3.48 (3.36, 3.61) | 2.02 (1.90, 2.14) |
| | PMR | 1.65 (1.61, 1.69) | 0.79 (0.75, 0.84) | 2.19 (2.13, 2.24) | 1.49 (1.43, 1.56) | 2.23 (2.16, 2.30) | 3.26 (2.78, 3.82) | 0.83 (0.73, 0.93) | 1.03 (1.02, 1.04) | 1.53 (1.50, 1.56) | 0.73 (0.71, 0.74) | 0.55 (0.53, 0.57) | 2.79 (2.72, 2.86) | 1.69 (1.66, 1.71) | 1.04 (1.01, 1.06) | 5.05 (4.92, 5.19) | 3.34 (3.18, 3.51) | 1.40 (1.27, 1.55) |
| | PM | 0.51 (0.47, 0.54) | 0.24 (0.21, 0.29) | 0.64 (0.60, 0.69) | 0.48 (0.42, 0.54) | 0.60 (0.54, 0.66) | 0.22 (0.09, 0.53) | 0.27 (0.20, 0.38) | 0.74 (0.72, 0.76) | 0.65 (0.62, 0.69) | 0.50 (0.48, 0.52) | 0.53 (0.51, 0.56) | 1.90 (1.81, 1.99) | 2.33 (2.29, 2.38) | 1.46 (1.41, 1.50) | 6.74 (6.54, 6.96) | 4.81 (4.52, 5.11) | 1.14 (0.96, 1.35) |
| | Neuro | 1.45 (1.40, 1.51) | 0.68 (0.61, 0.75) | 1.96 (1.88, 2.05) | 1.38 (1.27, 1.49) | 1.85 (1.75, 1.95) | 0.67 (0.39, 1.13) | 0.07 (0.04, 0.14) | 1.20 (1.18, 1.21) | 1.98 (1.93, 2.03) | 0.61 (0.58, 0.63) | 0.69 (0.66, 0.72) | 4.64 (4.51, 4.77) | 1.87 (1.83, 1.92) | 1.28 (1.23, 1.33) | 3.81 (3.64, 3.99) | 9.71 (9.29, 10.16) | 6.42 (5.96, 6.93) |
| | Neurologist | 1.11 (1.06, 1.16) | 1.05 (0.97, 1.15) | 1.28 (1.21, 1.35) | 1.16 (1.07, 1.27) | 1.19 (1.11, 1.28) | 1.47 (1.02, 2.11) | 0.21 (0.14, 0.31) | 0.99 (0.98, 1.01) | 0.75 (0.71, 0.79) | 0.74 (0.71, 0.77) | 0.79 (0.76, 0.82) | 3.02 (2.90, 3.13) | 1.17 (1.13, 1.22) | 0.90 (0.86, 0.94) | 2.39 (2.25, 2.53) | 2.53 (2.31, 2.76) | 2.07 (1.81, 2.36) |
| | Rheu | 0.87 (0.82, 0.92) | 1.05 (0.97, 1.15) | 0.90 (0.84, 0.96) | 0.85 (0.76, 0.94) | 0.88 (0.81, 0.96) | 1.44 (0.99, 2.12) | 0.17 (0.11, 0.27) | 1.05 (1.03, 1.06) | 1.54 (1.49, 1.59) | 0.90 (0.87, 0.94) | 0.57 (0.55, 0.60) | 1.51 (1.42, 1.60) | 1.05 (1.01, 1.09) | 0.90 (0.86, 0.95) | 1.92 (1.79, 2.05) | 1.57 (1.40, 1.77) | 1.23 (1.03, 1.46) |
| | MD (Oth) | 1.21 (1.17, 1.25) | 1.90 (1.81, 1.98) | 0.99 (0.94, 1.03) | 1.22 (1.15, 1.29) | 0.97 (0.92, 1.03) | 2.35 (1.92, 2.89) | 0.77 (0.67, 0.89) | 0.74 (0.72, 0.75) | 0.84 (0.82, 0.87) | 0.72 (0.70, 0.74) | 0.52 (0.50, 0.54) | 1.04 (0.99, 1.09) | 1.26 (1.23, 1.29) | 1.25 (1.22, 1.28) | 1.18 (1.11, 1.26) | 1.22 (1.11, 1.33) | 1.56 (1.40, 1.73) |
| Emergency/ Urgent Care | EM | 0.69 (0.66, 0.71) | 0.72 (0.68, 0.76) | 0.75 (0.71, 0.78) | 0.74 (0.70, 0.79) | 0.72 (0.69, 0.76) | 0.82 (0.62, 1.09) | 0.17 (0.13, 0.22) | 1.11 (1.10, 1.12) | 1.38 (1.35, 1.41) | 1.10 (1.08, 1.12) | 1.25 (1.23, 1.27) | 0.93 (0.89, 0.97) | 1.41 (1.38, 1.44) | 1.33 (1.30, 1.36) | 0.73 (0.69, 0.78) | 0.92 (0.85, 1.01) | 5.86 (5.57, 6.18) |
| | Rad | 0.27 (0.26, 0.29) | 0.09 (0.08, 0.11) | 0.38 (0.35, 0.40) | 0.22 (0.19, 0.25) | 0.34 (0.31, 0.37) | 0.28 (0.16, 0.49) | 0.03 (0.01, 0.06) | 1.41 (1.40, 1.42) | 3.12 (3.08, 3.16) | 0.07 (0.06, 0.07) | 0.05 (0.05, 0.06) | 2.33 (2.26, 2.41) | 0.48 (0.46, 0.50) | 0.07 (0.07, 0.08) | 0.63 (0.58, 0.68) | 0.61 (0.54, 0.69) | 5.17 (4.87, 5.50) |
| | UC | 0.65 (0.60, 0.70) | 0.72 (0.64, 0.82) | 0.67 (0.61, 0.74) | 0.63 (0.54, 0.72) | 0.68 (0.61, 0.77) | 0.87 (0.49, 1.53) | 0.31 (0.21, 0.46) | 0.95 (0.92, 0.97) | 1.03 (0.98, 1.09) | 0.85 (0.81, 0.89) | 1.16 (1.12, 1.20) | 0.48 (0.42, 0.54) | 0.75 (0.70, 0.79) | 0.77 (0.73, 0.82) | 0.52 (0.44, 0.61) | 0.53 (0.42, 0.67) | 0.59 (0.44, 0.80) |

PCP=Primary Care Provider, PA=Physician Assistant, DO=Doctor of Osteopathy, DC=Doctor of Chiropractic, PT=Physical Therapist, LAc=Licensed Acupuncturist, OS=Orthpedic Surgeon, PMR=Physical Medicine & Rehabilitation, PM=Pain Management, NS=Neurosurgeon, Neuro=Neurologist, Rheum=Rheumatologist, MD Oth=Other MD specialty, EM=Emergency Medicine, Rad=Radiologist, UC=Urgent Care

Cells in red are not different than the PCP reference (p=.05)
