## Supplement 5 - Non-surgical Timing for "Low back pain care pathways and costs: association with the type of initial contact health care provider. A retrospective cohort study"

### Supplement 5 - Low back pain # of days into episode when service initially provided by type of initial contact health care provider (HCP)

| All Non-Surgical Episodes Median (IQR) (Q1, Q3) | | First Line | | | | | | |
|---|---|---|---|---|---|---|---|---|
| | | Any | Manipulation - Chiropractic | Active Care | Passive Therapy | Manual Therapy | Acupuncture | Manipulation - Osteopathic |
| Primary Care | PCP-reference | 22 (65) (5, 70) | 44 (93) (12, 105) | 23 (63) (7, 70) | 32 (82) (8, 90) | 24 (62) (8, 70) | 36 (100) (8, 108) | 0 (0) (0, 0) |
| | Nurse | 21 (66) (4, 70) | 31 (83) (6, 89) | 21 (62) (6, 68) | 26 (81) (6, 87) | 23 (63) (6, 69) | 38 (141) (10, 151) | 1 (33) (0, 33) |
| | PA | 21 (58) (7, 65) | 35 (83) (8, 91) | 20 (53) (7, 60) | 33 (82) (9, 91) | 24 (57) (8, 65) | 40 (87) (12, 99) | 22 (62) (2, 64) |
| | DO | 0 (0) (0, 0) | 88 (100) (27, 128) | 14 (48) (0, 48) | 9 (130) (0, 130) | 34 (82) (8, 90) | 41 (119) (25, 144) | 0 (0) (0, 0) |
| | All | 21 (64) (5, 69) | 41 (91) (10, 101) | 22 (61) (7, 68) | 31 (82) (8, 90) | 24 (61) (8, 69) | 37 (100) (8, 108) | 0 (1) (0, 1) |
| Non-Prescriber | DC | 0 (0) (0, 0) | 0 (0) (0, 0) | 0 (4) (0, 4) | 0 (0) (0, 0) | 0 (6) (0, 6) | 8 (58) (0, 58) | 35 (120) (1, 121) |
| | PT | 0 (0) (0, 0) | 35 (64) (8, 72) | 0 (1) (0, 1) | 4 (23) (0, 23) | 0 (7) (0, 7) | 22 (80) (0, 80) | 58 (114) (22, 136) |
| | LAc | 0 (0) (0, 0) | 13 (51) (0, 51) | 1 (35) (0, 35) | 0 (0) (0, 0) | 0 (0) (0, 0) | 0 (0) (0, 0) | 1 (30) (0, 30) |
| | All | 0 (0) (0, 0) | 0 (0) (0, 0) | 0 (4) (0, 4) | 0 (1) (0, 1) | 0 (7) (0, 7) | 0 (0) (0, 0) | 38 (120) (1, 121) |
| Specialist | OS | 15 (38) (6, 44) | 53 (105) (16, 121) | 14 (34) (6, 40) | 23 (62) (7, 69) | 17 (38) (7, 45) | 61 (108) (21, 129) | 0 (40) (0, 40) |
| | PMR | 13 (37) (3, 40) | 42 (119) (7, 126) | 14 (35) (4, 39) | 21 (66) (5, 71) | 16 (39) (5, 44) | 22 (109) (0, 109) | 0 (21) (0, 21) |
| | PM | 11 (50) (0, 50) | 37 (136) (4, 140) | 11 (48) (0, 48) | 20 (86) (0, 86) | 12 (49) (0, 49) | 61 (62) (0, 62) | 0 (6) (0, 6) |
| | NS | 27 (63) (8, 71) | 64 (126) (13, 139) | 27 (54) (8, 62) | 37 (78) (13, 91) | 29 (56) (11, 68) | 148 (111) (114, 225) | 157 (148) (80, 228) |
| | Neuro | 39 (92) (12, 104) | 60 (99) (23, 122) | 39 (93) (12, 105) | 49 (116) (15, 132) | 36 (94) (13, 108) | 52 (77) (32, 109) | 42 (83) (0, 83) |
| | Rheu | 54 (112) (19, 131) | 56 (114) (22, 136) | 61 (116) (21, 136) | 66 (139) (29, 168) | 68 (121) (22, 143) | 62 (142) (18, 159) | 58 (151) (27, 178) |
| | MD (Oth) | 51 (97) (15, 112) | 54 (91) (19, 110) | 63 (101) (23, 124) | 66 (102) (28, 130) | 63 (100) (24, 124) | 68 (117) (22, 140) | 0 (25) (0, 25) |
| | All | 19 (55) (6, 61) | 53 (104) (16, 120) | 18 (48) (6, 54) | 31 (81) (9, 90) | 21 (51) (7, 58) | 50 (118) (10, 128) | 0 (34) (0, 34) |
| Emergency/ Urgent Care | EM | 19 (42) (6, 48) | 18 (59) (4, 63) | 20 (40) (8, 48) | 23 (51) (7, 58) | 23 (40) (10, 50) | 28 (103) (10, 113) | 8 (39) (2, 41) |
| | Rad | 18 (31) (6, 37) | 27 (90) (4, 94) | 19 (30) (6, 36) | 23 (46) (10, 56) | 20 (35) (7, 42) | 84 (100) (49, 149) | 18 (30) (9, 38) |
| | UC | 17 (43) (5, 48) | 16 (60) (5, 65) | 21 (46) (10, 56) | 21 (58) (7, 65) | 20 (42) (9, 51) | 26 (25) (16, 40) | 0 (6) (0, 6) |
| | All | 18 (39) (6, 45) | 18 (61) (4, 65) | 20 (37) (8, 45) | 23 (50) (8, 58) | 21 (40) (9, 49) | 34 (104) (14, 118) | 7 (31) (0, 31) |

| All Non-Surgical Episodes Median (IQR) (Q1, Q3) | | Second Line | | | | | Third Line | | | |
|---|---|---|---|---|---|---|---|---|---|---|
| | | Any | Imaging - Radiography | Rx - NSAID | Rx - Skeletal Muscle Relaxant | Imaging - MRI | Any | Rx-Opioid | Spinal Injection | Imaging-CT |
| Primary Care | PCP-reference | 0 (1) (0, 1) | 1 (38) (0, 38) | 0 (8) (0, 8) | 0 (1) (0, 1) | 35 (100) (10, 110) | 0 (42) (0, 42) | 0 (30) (0, 30) | 60 (126) (14, 140) | 29 (119) (1, 120) |
| | Nurse | 0 (0) (0, 0) | 0 (33) (0, 33) | 0 (4) (0, 4) | 0 (0) (0, 0) | 34 (96) (10, 106) | 1 (47) (0, 47) | 0 (42) (0, 42) | 41 (108) (7, 115) | 17 (108) (0, 108) |
| | PA | 0 (0) (0, 0) | 0 (12) (0, 12) | 0 (3) (0, 3) | 0 (0) (0, 0) | 21 (69) (7, 76) | 1 (36) (0, 36) | 0 (31) (0, 31) | 35 (88) (7, 95) | 8 (74) (0, 74) |
| | DO | 1 (47) (0, 47) | 8 (87) (0, 87) | 14 (61) (0, 61) | 2 (42) (0, 42) | 30 (80) (10, 90) | 27 (90) (2, 92) | 24 (96) (0, 96) | 50 (77) (14, 91) | 80 (182) (2, 185) |
| | All | 0 (0) (0, 0) | 0 (34) (0, 34) | 0 (7) (0, 7) | 0 (1) (0, 1) | 33 (96) (9, 105) | 0 (42) (0, 42) | 0 (32) (0, 32) | 53 (119) (12, 131) | 23 (112) (0, 112) |
| Non-Prescriber | DC | 0 (51) (0, 51) | 0 (3) (0, 3) | 76 (129) (25, 154) | 47 (120) (9, 129) | 57 (114) (19, 133) | 65 (122) (19, 141) | 70 (124) (22, 146) | 64 (130) (16, 146) | 52 (123) (4, 127) |
| | PT | 15 (57) (0, 57) | 10 (55) (0, 55) | 48 (112) (14, 126) | 30 (81) (4, 85) | 41 (78) (8, 86) | 45 (91) (12, 103) | 45 (112) (9, 121) | 57 (84) (22, 106) | 61 (116) (7, 123) |
| | LAc | 44 (87) (8, 95) | 39 (72) (6, 78) | 56 (127) (20, 146) | 43 (85) (3, 88) | 63 (97) (31, 128) | 51 (122) (14, 136) | 51 (138) (18, 156) | 55 (117) (8, 125) | 91 (208) (32, 239) |
| | All | 1 (52) (0, 52) | 0 (6) (0, 6) | 73 (129) (24, 153) | 45 (118) (8, 126) | 53 (110) (17, 127) | 63 (120) (18, 138) | 68 (124) (21, 145) | 62 (121) (17, 138) | 54 (123) (4, 128) |
| Specialist | OS | 0 (0) (0, 0) | 0 (0) (0, 0) | 0 (28) (0, 28) | 0 (37) (0, 37) | 10 (34) (2, 36) | 13 (60) (0, 60) | 7 (63) (0, 63) | 31 (72) (6, 78) | 30 (102) (3, 105) |
| | PMR | 0 (12) (0, 12) | 0 (6) (0, 6) | 6 (73) (0, 73) | 0 (48) (0, 48) | 11 (46) (2, 48) | 7 (36) (0, 36) | 3 (45) (0, 45) | 17 (53) (0, 53) | 49 (124) (7, 130) |
| | PM | 0 (27) (0, 27) | 0 (39) (0, 39) | 5 (73) (0, 73) | 1 (50) (0, 50) | 13 (49) (3, 52) | 0 (22) (0, 22) | 0 (17) (0, 17) | 14 (49) (0, 49) | 65 (157) (12, 169) |
| | NS | 0 (12) (0, 12) | 0 (23) (0, 23) | 28 (117) (0, 117) | 14 (84) (0, 84) | 7 (26) (0, 26) | 20 (64) (0, 64) | 17 (84) (0, 84) | 42 (74) (18, 92) | 18 (93) (0, 93) |
| | Neuro | 0 (27) (0, 27) | 40 (116) (3, 119) | 11 (81) (0, 81) | 0 (41) (0, 41) | 14 (61) (1, 62) | 18 (86) (0, 86) | 13 (88) (0, 88) | 45 (110) (10, 119) | 43 (135) (6, 141) |
| | Rheu | 0 (15) (0, 15) | 0 (37) (0, 37) | 0 (40) (0, 40) | 1 (70) (0, 70) | 49 (127) (14, 141) | 17 (95) (0, 95) | 14 (85) (0, 85) | 55 (141) (2, 143) | 69 (134) (17, 151) |
| | MD (Oth) | 4 (71) (0, 71) | 34 (109) (0, 109) | 2 (83) (0, 83) | 17 (95) (0, 95) | 64 (131) (15, 146) | 0 (40) (0, 40) | 0 (29) (0, 29) | 77 (135) (22, 157) | 33 (116) (0, 116) |
| | All | 0 (6) (0, 6) | 0 (3) (0, 3) | 0 (53) (0, 53) | 0 (55) (0, 55) | 12 (47) (2, 49) | 7 (49) (0, 49) | 3 (51) (0, 51) | 26 (75) (1, 76) | 33 (116) (1, 117) |
| Emergency/ Urgent Care | EM | 0 (0) (0, 0) | 0 (1) (0, 1) | 0 (1) (0, 1) | 0 (1) (0, 1) | 12 (41) (1, 42) | 0 (1) (0, 1) | 0 (1) (0, 1) | 28 (76) (4, 80) | 0 (0) (0, 0) |
| | Rad | 0 (0) (0, 0) | 0 (0) (0, 0) | 13 (64) (0, 64) | 1 (38) (0, 38) | 0 (0) (0, 0) | 0 (8) (0, 8) | 12 (55) (0, 55) | 29 (59) (10, 69) | 0 (0) (0, 0) |
| | UC | 0 (0) (0, 0) | 0 (1) (0, 1) | 0 (0) (0, 0) | 0 (0) (0, 0) | 30 (73) (9, 82) | 0 (14) (0, 14) | 0 (6) (0, 6) | 61 (137) (7, 144) | 0 (40) (0, 40) |
| | All | 0 (0) (0, 0) | 0 (0) (0, 0) | 0 (1) (0, 1) | 0 (1) (0, 1) | 0 (15) (0, 15) | 0 (1) (0, 1) | 0 (2) (0, 2) | 30 (72) (6, 78) | 0 (0) (0, 0) |

PCP=Primary Care Provider, PA=Physician Assistant, DO=Doctor of Osteopathy, DC=Doctor of Chiropractic, PT=Physical Therapist, LAc=Licensed Acupuncturist, OS=Orthpedic Surgeon, PMR=Physical Medicine & Rehabilitation, PM=Pain Management, NS=Neurosurgeon, Neuro=Neurologist, Rheum=Rheumatologist, MD Oth=Other MD specialty, EM=Emergency Medicine, Rad=Radiologist, UC=Urgent Care, IQR=Interquartile Range

Cells with red text denote that the effect of provider type on service usage was found not to be significantly different from that of PCP-reference (Mann-Whitney U $p > 0.001$)
Cells with black text denote that the effect of provider type on service usage was found to be significantly different from that of PCP-reference (Mann-Whitney U $p < 0.001$)
