## Supplement 6 - Pooled Timing for "Low back pain care pathways and costs: association with the type of initial contact health care provider. A retrospective cohort study"

**Supplement 6 - Low back pain # of days into episode when service initially provided by type of initial contact health care provider (HCP)**

| Combined Surgical and Non-Surgical (Pooled) Episodes Median (IQR)(Q1, Q3) | | First Line | | | | | | |
|---|---|---|---|---|---|---|---|---|
| | | Any | Manipulation - Chiropractic | Active Care | Passive Therapy | Manual Therapy | Acupuncture | Manipulation - Osteopathic |
| Primary Care | PCP-reference | 23 (70) (5, 75) | 45 (95) (12, 107) | 26 (70) (8, 78) | 34 (87) (9, 96) | 27 (69) (8, 77) | 37 (103) (8, 111) | 0 (0) (0, 0) |
| | Nurse | 23 (70) (5, 75) | 32 (84) (6, 90) | 23 (70) (6, 76) | 28 (86) (6, 92) | 25 (71) (7, 78) | 45 (137) (13, 150) | 2 (36) (0, 36) |
| | PA | 23 (63) (7, 70) | 36 (86) (8, 94) | 22 (61) (7, 68) | 35 (86) (10, 96) | 26 (66) (8, 74) | 44 (88) (14, 101) | 23 (67) (3, 70) |
| | DO | 0 (0) (0, 0) | 88 (100) (27, 128) | 14 (52) (0, 52) | 9 (130) (0, 130) | 34 (82) (8, 90) | 42 (154) (30, 184) | 0 (0) (0, 0) |
| | All | 23 (69) (5, 74) | 42 (93) (10, 103) | 25 (70) (7, 77) | 33 (87) (8, 95) | 27 (69) (8, 77) | 39 (104) (9, 113) | 0 (1) (0, 1) |
| Non-Prescriber | DC | 0 (0) (0, 0) | 0 (0) (0, 0) | 0 (4) (0, 4) | 0 (1) (0, 1) | 0 (7) (0, 7) | 8 (59) (0, 59) | 38 (123) (1, 124) |
| | PT | 0 (0) (0, 0) | 36 (76) (9, 84) | 0 (2) (0, 2) | 5 (24) (0, 24) | 0 (8) (0, 8) | 22 (84) (0, 85) | 58 (121) (20, 141) |
| | LAc | 0 (0) (0, 0) | 14 (52) (0, 52) | 2 (35) (0, 35) | 0 (0) (0, 0) | 0 (0) (0, 0) | 0 (0) (0, 0) | 1 (30) (0, 30) |
| | All | 0 (0) (0, 0) | 0 (0) (0, 0) | 0 (4) (0, 4) | 0 (1) (0, 1) | 0 (7) (0, 7) | 0 (0) (0, 0) | 40 (122) (2, 124) |
| Specialist | OS | 17 (48) (6, 54) | 54 (107) (17, 124) | 17 (44) (6, 50) | 27 (69) (8, 77) | 20 (49) (7, 56) | 64 (108) (22, 130) | 0 (43) (0, 43) |
| | PMR | 14 (45) (4, 49) | 48 (125) (8, 133) | 15 (43) (5, 48) | 25 (78) (6, 84) | 19 (47) (6, 53) | 26 (114) (0, 114) | 0 (30) (0, 30) |
| | PM | 15 (69) (0, 69) | 43 (132) (8, 140) | 15 (65) (1, 66) | 28 (110) (2, 112) | 17 (66) (0, 66) | 61 (62) (0, 62) | 0 (6) (0, 6) |
| | NS | 41 (87) (12, 99) | 71 (135) (17, 152) | 41 (85) (13, 98) | 52 (96) (19, 116) | 44 (91) (15, 106) | 138 (144) (64, 208) | 157 (101) (117, 218) |
| | Neuro | 42 (98) (13, 110) | 62 (102) (24, 126) | 43 (100) (13, 113) | 52 (121) (16, 137) | 40 (103) (14, 117) | 58 (79) (32, 111) | 37 (82) (0, 82) |
| | Rheu | 57 (117) (21, 138) | 57 (118) (22, 140) | 63 (118) (23, 141) | 70 (144) (29, 173) | 74 (122) (27, 149) | 51 (122) (20, 142) | 104 (135) (36, 171) |
| | MD (Oth) | 53 (100) (15, 115) | 54 (94) (19, 113) | 68 (107) (25, 132) | 68 (105) (28, 133) | 67 (106) (26, 132) | 72 (121) (25, 146) | 0 (31) (0, 31) |
| | All | 22 (65) (7, 72) | 55 (107) (17, 124) | 21 (62) (7, 69) | 35 (89) (10, 99) | 25 (65) (8, 73) | 52 (119) (11, 130) | 0 (42) (0, 42) |
| Emergency/ Urgent Care | EM | 19 (44) (6, 50) | 18 (60) (4, 64) | 21 (43) (8, 51) | 24 (52) (8, 60) | 24 (43) (10, 53) | 27 (104) (10, 114) | 9 (66) (2, 68) |
| | Rad | 20 (36) (6, 42) | 29 (91) (4, 95) | 20 (35) (7, 42) | 26 (50) (11, 60) | 21 (41) (7, 48) | 84 (100) (49, 149) | 18 (30) (9, 38) |
| | UC | 18 (45) (6, 51) | 16 (64) (5, 68) | 23 (54) (11, 65) | 24 (59) (7, 66) | 21 (48) (10, 58) | 22 (22) (15, 36) | 0 (6) (0, 6) |
| | All | 19 (43) (6, 49) | 18 (64) (4, 68) | 21 (41) (8, 49) | 24 (52) (8, 60) | 23 (43) (9, 52) | 32 (106) (12, 118) | 8 (41) (0, 41) |

| Combined Surgical and Non-Surgical (Pooled) Episodes Median (IQR)(Q1, Q3) | | Second Line | | | | | Third Line | | | | |
|---|---|---|---|---|---|---|---|---|---|---|---|
| | | Any | Imaging - Radiography | Rx - NSAID | Rx - Skeletal Muscle Relaxant | Imaging - MRI | Any | Rx-Opioid | Spinal Injection | Spinal Surgery | Imaging-CT |
| Primary Care | PCP-reference | 0 (1) (0, 1) | 1 (45) (0, 45) | 0 (11) (0, 11) | 0 (2) (0, 2) | 35 (101) (10, 111) | 0 (49) (0, 49) | 0 (38) (0, 38) | 62 (119) (20, 139) | 64 (107) (26, 133) | 41 (133) (2, 135) |
| | Nurse | 0 (0) (0, 0) | 0 (40) (0, 40) | 0 (7) (0, 7) | 0 (0) (0, 0) | 36 (98) (10, 108) | 3 (53) (0, 53) | 1 (50) (0, 50) | 47 (109) (11, 120) | 55 (108) (18, 126) | 28 (129) (0, 129) |
| | PA | 0 (0) (0, 0) | 0 (17) (0, 17) | 0 (7) (0, 7) | 0 (0) (0, 0) | 21 (73) (7, 80) | 2 (43) (0, 43) | 1 (40) (0, 40) | 39 (90) (11, 102) | 44 (90) (16, 106) | 16 (101) (0, 101) |
| | DO | 1 (50) (0, 50) | 8 (85) (0, 85) | 14 (62) (0, 62) | 3 (51) (0, 51) | 35 (80) (12, 92) | 28 (92) (3, 94) | 29 (96) (0, 97) | 50 (82) (14, 96) | 125 (152) (54, 206) | 11 (172) (3, 175) |
| | All | 0 (0) (0, 0) | 1 (41) (0, 41) | 0 (10) (0, 10) | 0 (1) (0, 1) | 34 (98) (9, 107) | 1 (49) (0, 49) | 0 (40) (0, 40) | 56 (115) (16, 131) | 60 (105) (23, 128) | 35 (129) (1, 130) |
| Non-Prescriber | DC | 0 (52) (0, 52) | 0 (6) (0, 6) | 75 (129) (24, 153) | 48 (122) (9, 131) | 57 (112) (20, 132) | 64 (121) (19, 140) | 70 (124) (22, 146) | 68 (126) (20, 146) | 76 (116) (32, 148) | 62 (134) (7, 141) |
| | PT | 16 (57) (0, 57) | 15 (58) (0, 58) | 48 (114) (14, 128) | 35 (92) (6, 98) | 40 (78) (9, 87) | 44 (85) (12, 97) | 52 (111) (12, 123) | 56 (83) (21, 104) | 53 (73) (21, 94) | 70 (131) (24, 155) |
| | LAc | 43 (86) (8, 94) | 39 (76) (7, 83) | 52 (124) (19, 143) | 47 (100) (5, 105) | 61 (94) (28, 122) | 52 (119) (15, 134) | 55 (137) (20, 156) | 53 (113) (12, 125) | 70 (120) (29, 148) | 94 (190) (32, 222) |
| | All | 1 (54) (0, 54) | 0 (8) (0, 8) | 72 (129) (23, 152) | 47 (120) (9, 129) | 53 (109) (17, 126) | 62 (118) (18, 136) | 68 (124) (21, 145) | 64 (119) (20, 139) | 70 (112) (29, 141) | 63 (137) (8, 145) |
| Specialist | OS | 0 (0) (0, 0) | 0 (0) (0, 0) | 0 (37) (0, 37) | 1 (59) (0, 59) | 10 (37) (3, 40) | 16 (58) (0, 58) | 15 (77) (0, 77) | 32 (69) (8, 77) | 34 (63) (11, 74) | 45 (118) (8, 126) |
| | PMR | 0 (14) (0, 14) | 0 (16) (0, 16) | 9 (80) (0, 80) | 4 (68) (0, 68) | 13 (52) (3, 55) | 8 (37) (0, 37) | 7 (63) (0, 63) | 18 (56) (0, 56) | 22 (66) (1, 67) | 68 (148) (13, 161) |
| | PM | 1 (29) (0, 29) | 4 (62) (0, 62) | 7 (80) (0, 80) | 2 (63) (0, 63) | 13 (60) (2, 62) | 0 (22) (0, 22) | 0 (23) (0, 23) | 14 (46) (0, 46) | 11 (39) (0, 39) | 75 (178) (15, 193) |
| | NS | 0 (16) (0, 16) | 5 (49) (0, 49) | 35 (126) (1, 127) | 30 (95) (1, 96) | 8 (36) (0, 36) | 19 (55) (0, 55) | 29 (85) (2, 87) | 41 (82) (16, 98) | 30 (65) (5, 70) | 28 (102) (4, 106) |
| | Neuro | 0 (30) (0, 30) | 45 (118) (6, 124) | 14 (88) (0, 88) | 0 (51) (0, 51) | 14 (67) (2, 69) | 21 (87) (0, 87) | 19 (95) (0, 95) | 47 (110) (14, 124) | 50 (106) (15, 122) | 58 (140) (7, 146) |
| | Rheu | 0 (17) (0, 17) | 0 (46) (0, 46) | 0 (42) (0, 42) | 5 (78) (0, 78) | 51 (132) (14, 146) | 19 (96) (0, 96) | 18 (92) (0, 92) | 61 (136) (9, 146) | 68 (123) (21, 144) | 84 (134) (20, 154) |
| | MD (Oth) | 5 (73) (0, 73) | 38 (114) (0, 114) | 4 (88) (0, 88) | 20 (99) (0, 99) | 69 (133) (16, 149) | 0 (48) (0, 48) | 0 (37) (0, 37) | 77 (132) (22, 154) | 66 (131) (17, 148) | 45 (135) (0, 135) |
| | All | 0 (8) (0, 8) | 0 (13) (0, 13) | 2 (62) (0, 62) | 5 (71) (0, 71) | 13 (52) (2, 54) | 10 (50) (0, 50) | 8 (66) (0, 66) | 27 (73) (4, 77) | 30 (69) (7, 76) | 47 (127) (7, 134) |
| Emergency/ Urgent Care | EM | 0 (0) (0, 0) | 0 (3) (0, 3) | 0 (1) (0, 1) | 0 (1) (0, 1) | 12 (40) (2, 42) | 0 (1) (0, 1) | 0 (2) (0, 2) | 31 (74) (7, 82) | 26 (70) (7, 76) | 0 (0) (0, 0) |
| | Rad | 0 (0) (0, 0) | 0 (0) (0, 0) | 14 (74) (0, 74) | 2 (58) (0, 58) | 0 (0) (0, 0) | 0 (16) (0, 16) | 18 (76) (0, 76) | 27 (55) (8, 62) | 22 (49) (3, 52) | 0 (0) (0, 0) |
| | UC | 0 (0) (0, 0) | 0 (3) (0, 3) | 0 (0) (0, 0) | 0 (0) (0, 0) | 26 (66) (9, 75) | 0 (20) (0, 20) | 0 (10) (0, 10) | 50 (119) (13, 132) | 36 (74) (19, 93) | 0 (45) (0, 45) |
| | All | 0 (0) (0, 0) | 0 (0) (0, 0) | 0 (2) (0, 2) | 0 (1) (0, 1) | 0 (18) (0, 18) | 0 (2) (0, 2) | 0 (3) (0, 3) | 30 (69) (8, 77) | 26 (65) (7, 72) | 0 (0) (0, 0) |

PCP=Primary Care Provider, PA=Physician Assistant, DO=Doctor of Osteopathy, DC=Doctor of Chiropractic, PT=Physical Therapist, LAc=Licensed Acupuncturist, OS=Orthpedic Surgeon, PMR=Physical Medicine & Rehabilitation, PM=Pain Management, NS=Neurosurgeon, Neuro=Neurologist, Rheum=Rheumatologist, MD Oth=Other MD specialty, EM=Emergency Medicine, Rad=Radiologist, UC=Urgent Care, IQR=Interquartile Range

Cells with red text denote that the effect of provider type on service usage was found not to be significantly different from that of PCP-reference (Mann-Whitney U $p > 0.001$)
Cells with black text denote that the effect of provider type on service usage was found to be significantly different from that of PCP-reference (Mann-Whitney U $p < 0.001$)
