## Supplement 7 - Pooled Cost for "Low back pain care pathways and costs: association with the type of initial contact health care provider. A retrospective cohort study"

| Supplement 7 - Low back pain total episode cost by episode sequence cohort | | | | | | |
|---|---|---|---|---|---|---|
| Median (IQR) (Q1, Q3) | | Combined Surgical and Non-surgical (Pooled) Episodes | | | | |
| | | All | Single | Multiple - First | Multiple Second | Multiple - Third+ |
| Primary Care | PCP-reference | $147 (408) (52, 460) | $149 (407) (57, 464) | $153 (452) (52, 504) | $132 (413) (31, 444) | $95 (222) (24, 246) |
| | Nurse | $152 (502) (38, 539) | $155 (496) (45, 540) | $160 (567) (38, 604) | $132 (499) (21, 520) | $103 (268) (23, 290) |
| | PA | $203 (688) (62, 750) | $206 (664) (70, 735) | $216 (860) (60, 920) | $178 (719) (30, 749) | $113 (356) (19, 375) |
| | DO | $224 (528) (92, 619) | $234 (511) (100, 611) | $219 (640) (88, 728) | $195 (554) (92, 646) | $114 (165) (63, 228) |
| | All | $152 (443) (51, 494) | $155 (440) (57, 497) | $158 (496) (51, 547) | $136 (448) (29, 477) | $97 (235) (23, 258) |
| Non-Prescriber | DC | $165 (336) (66, 402) | $190 (381) (81, 461) | $162 (302) (68, 370) | $135 (274) (60, 334) | $90 (148) (50, 198) |
| | PT | $769 (1385) (355, 1740) | $749 (1345) (347, 1693) | $1019 (2060) (448, 2508) | $760 (1230) (348, 1578) | $560 (724) (348, 1072) |
| | LAc | $365 (645) (157, 802) | $357 (616) (146, 763) | $386 (658) (184, 842) | $406 (734) (186, 919) | $368 (569) (156, 725) |
| | All | $180 (365) (70, 435) | $200 (409) (90, 499) | $165 (325) (70, 395) | $142 (300) (60, 360) | $97 (150) (52, 202) |
| Specialist | OS | $455 (1378) (169, 1547) | $444 (1305) (173, 1478) | $561 (1660) (180, 1840) | $426 (1493) (141, 1635) | $279 (1109) (112, 1221) |
| | PMR | $800 (2012) (256, 2268) | $796 (2003) (262, 2265) | $898 (2073) (270, 2344) | $788 (2031) (231, 2262) | $427 (1409) (133, 1542) |
| | PM | $713 (1919) (212, 2130) | $787 (2055) (222, 2277) | $639 (1759) (214, 1973) | $612 (1627) (184, 1811) | $313 (1110) (163, 1272) |
| | NS | $1512 (5160) (378, 5538) | $1607 (5482) (406, 5889) | $1605 (5407) (397, 5804) | $1203 (3909) (307, 4215) | $722 (2698) (215, 2913) |
| | Neuro | $540 (1593) (152, 1745) | $603 (1707) (182, 1889) | $503 (1541) (153, 1694) | $389 (1203) (110, 1313) | $143 (562) (30, 593) |
| | Rheu | $339 (1025) (106, 1131) | $356 (1101) (106, 1207) | $368 (1230) (125, 1355) | $297 (769) (99, 869) | $251 (528) (74, 602) |
| | MD (Oth) | $142 (599) (22, 621) | $155 (679) (24, 703) | $165 (653) (25, 678) | $97 (370) (16, 386) | $80 (243) (17, 260) |
| | All | $492 (1592) (154, 1746) | $509 (1595) (163, 1758) | $557 (1789) (165, 1954) | $405 (1478) (119, 1596) | $253 (952) (84, 1036) |
| Emergency/ Urgent Care | EM | $590 (1451) (172, 1622) | $601 (1434) (182, 1616) | $636 (1592) (168, 1760) | $456 (1489) (78, 1567) | $325 (1260) (81, 1341) |
| | Rad | $192 (572) (56, 628) | $183 (524) (55, 579) | $265 (925) (70, 994) | $206 (740) (55, 794) | $177 (668) (63, 731) |
| | UC | $192 (257) (106, 363) | $191 (248) (110, 358) | $221 (293) (116, 410) | $179 (317) (63, 380) | $205 (358) (33, 391) |
| | All | $317 (1026) (95, 1122) | $316 (1002) (98, 1100) | $385 (1232) (110, 1342) | $273 (1042) (61, 1104) | $251 (847) (66, 913) |

PCP=Primary Care Provider, PA=Physician Assistant, DO=Doctor of Osteopathy, DC=Doctor of Chiropractic, PT=Physical Therapist, LAc=Licensed Acupuncturist, OS=Orthpedic Surgeon, PMR=Physical Medicine & Rehabilitation, PM=Pain Management, NS=Neurosurgeon, Neuro=Neurologist, Rheum=Rheumatologist, MD Oth=Other MD specialty, EM=Emergency Medicine, Rad=Radiologist, UC=Urgent Care, IQR=Interquartile Range

Cells with red text denote that the effect of provider type on service usage was found not to be significantly different from that of PCP-reference (Mann-Whitney U p > 0.001)

Cells with black text denote that the effect of provider type on service usage was found to be significantly different from that of PCP-reference (Mann-Whitney U p < 0.001)
