## Supplement - Care Pathways for "Low back pain care pathways and costs: association with the type of initial contact health care provider. A retrospective cohort study"

**Supplement - Care Pathways Page 1**

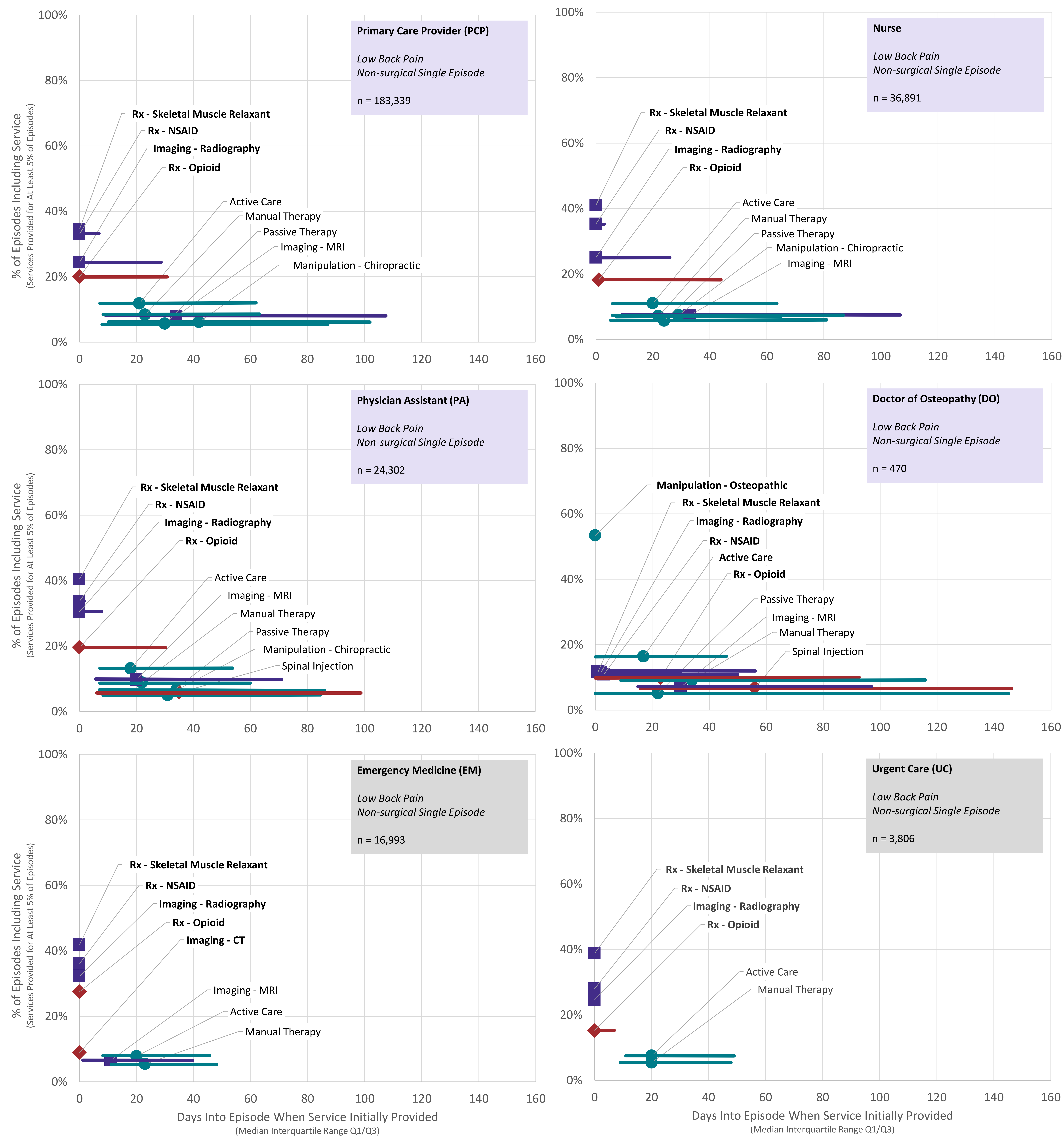


**Figure 3 Supplement.** Rate and timing of use of health care services for individuals with low back pain initially contacting a primary care provider, nurse, physician assistant, doctor of osteopathy, emergency medicine, and urgent care health care providers - non-surgical single episode cohort

**Supplement - Care Pathways Page 2**

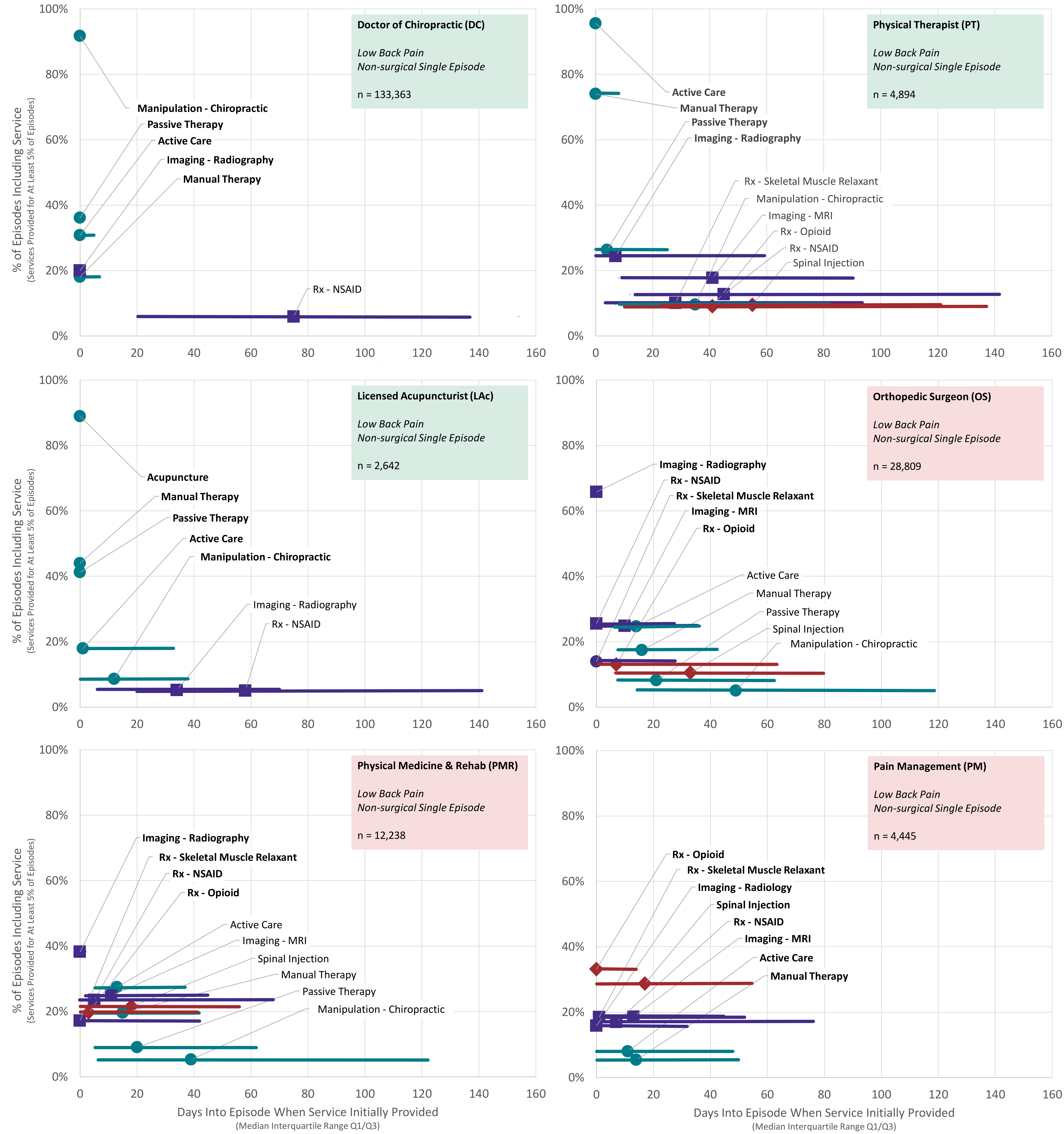


**Figure 3 Supplement.** Rate and timing of use of health care services for individuals with low back pain initially contacting a doctor of chiropractic, physical therapist, licensed acupuncturist assistant, orthopedic surgeon, physical medicine & rehabilitation, and pain management health care providers - non-surgical single episode cohort
