## Supplementary figures and images for "Low back pain care pathways and costs: association with the type of initial contact health care provider. A retrospective cohort study"

### Supplement - Cohort

Supplement – Cohort Description and Guide to Tables and Supplements

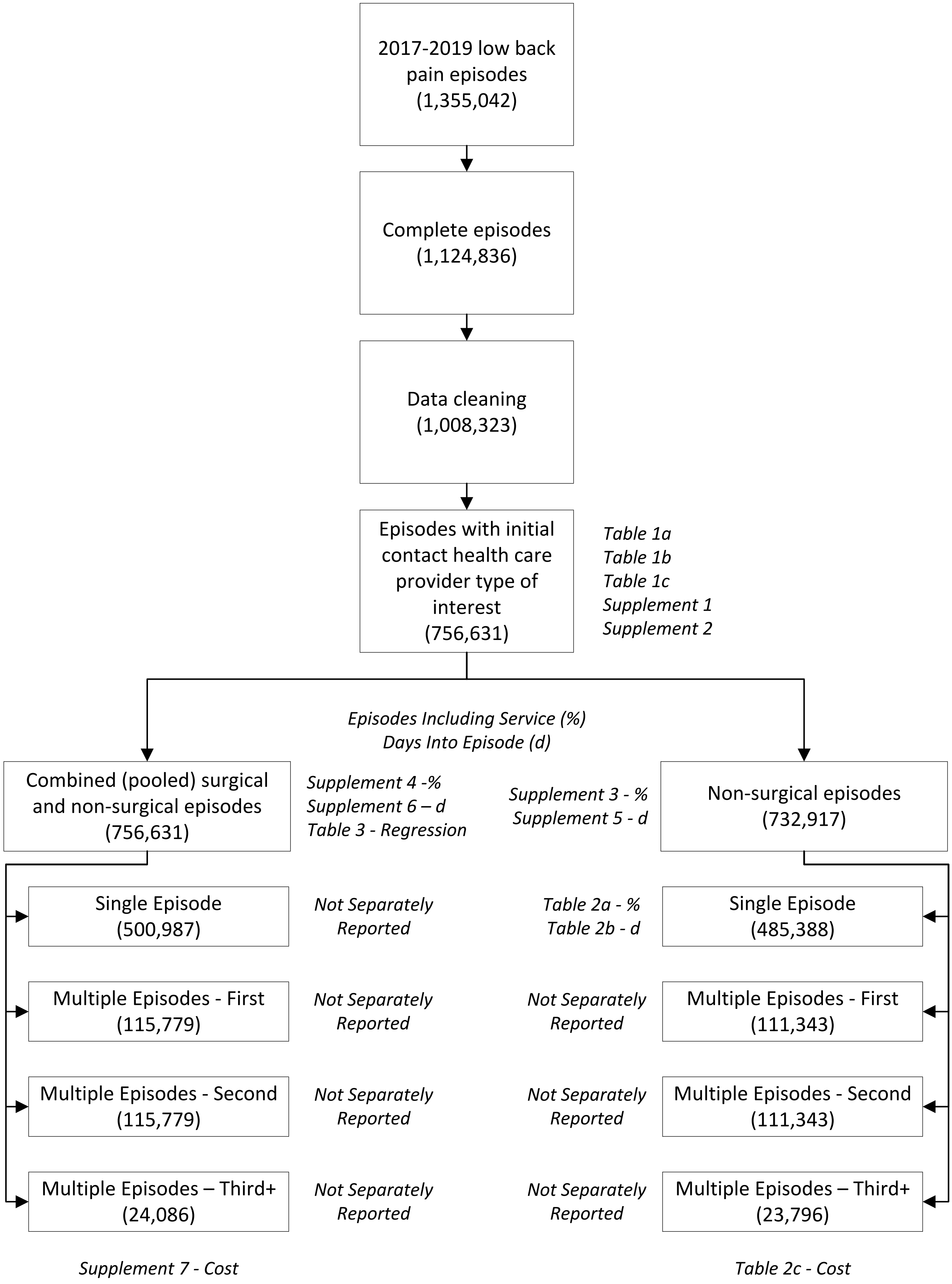
